# Supplementation with Arabinoxylan Dietary Fiber at Low Doses Produces Behavioral, Metabolic, and Gut Microbial Changes in Healthy, Overweight Adults: A Randomized Placebo-Controlled Trial

**DOI:** 10.64898/2026.06.08.26354814

**Authors:** Ainsley C. Arreza, Jun Wang, Stephanie-Anne Girard, Kelly A. Foley, Joshua Baisley, Stephanie Recker, Ambreen Atif, Hannah S. Ackermann, Andrew Richard

## Abstract

**Background:** Dietary fiber comprises a heterogeneous group of compounds with distinct physicochemical properties and biological effects. As such, functional outcomes observed for one fiber cannot be generalized to others. Some fermentable fibers, such as arabinoxylan, may exert biologically selective effects across multiple physiological domains, highlighting the need to evaluate individual ingredients for their domain-specific activity in controlled human studies.

**Methods:** In this randomized, double-blind, parallel, 3-arm, placebo-controlled trial, healthy, overweight adults were assigned to consume one of two low doses of an arabinoxylan dietary fiber (3.5g or 5g) or placebo over the intervention period. Self-reported appetite sensations were assessed as the primary outcome using validated visual analogue scales. Secondary and exploratory endpoints included lipid parameters, gastrointestinal outcomes, mood-related measures, and gut microbiota composition and fermentation-derived metabolites. Analyses were conducted in the full analysis set and a high-compliance population to assess responses under sustained intake conditions, as per the intended dosing regimen.

**Results:** The primary endpoint of appetite sensations did not differ between either arabinoxylan group and placebo. In contrast, evidence of microbial fermentation and selective microbiota engagement was observed. These responses occurred alongside consistent and favorable changes in lipid parameters under conditions of sustained intake, including reductions in low-density lipoprotein cholesterol and triglycerides. Additional outcomes, including gastrointestinal symptoms and mood, demonstrated domain-specific responses.

**Conclusion:** This study demonstrates that supplementation with low doses of arabinoxylan dietary fiber elicit biologically selective, domain-specific effects across metabolic, microbial, gastrointestinal, and behavioral outcomes, particularly under conditions of sustained intake. These responses occurred independently of changes in appetite sensation, indicating that functional effects were not mediated through appetite-related pathways. Collectively, the findings highlight the ingredient’s biological versatility and contextual responsiveness across physiological systems, and suggest its prebiotic potential through alignment with ISAPP’s definition of a prebiotic, supporting further investigation of specific mechanistic pathways.

**Clinical trial registration:** https://clinicaltrials.gov/study/NCT06884449, identifier: NCT06884449

## 1 Introduction

Dietary fiber is widely recognized for its role in digestive and metabolic health; however, it encompasses a broad range of compounds with distinct physicochemical properties and biological behaviors.(1) Consequently, health effects observed for one fiber cannot be assumed for others, underscoring the importance of evaluating individual fiber ingredients based on their specific characteristics.

The investigational product evaluated in the current study, Arrabina®, is a soluble, arabinoxylan-based dietary fiber. The arabinoxylan naturally present in Arrabina® is a non-digestible, complex carbohydrate composed of polymers containing xylose, arabinose, bound-polyphenols, and variable uronic acid residues.(2) Variation in polymer backbone length, side-chain structure, and the presence of bound phenolic compounds collectively contribute to the structural heterogeneity of arabinoxylan. These compositional features shape its physicochemical properties, fermentability, and interactions within the GI environment, including the release and subsequent metabolism of bound phenolic compounds during microbial fermentation. As a result, arabinoxylan exhibits variable biological engagement across intake levels, influencing its interactions with digestive processes and the gut microbiota.(3) For example, interaction of complex fibers with the gut microbiota through microbial utilization can result in the production of fermentation-related metabolites, including short-chain fatty acids (SCFAs).(4) In parallel, fiber structure and behavior within the gut may influence digestive dynamics and host physiological responses through mechanisms not exclusively dependent on microbial fermentation.(5,6) Together, these pathways represent overlapping and potentially complementary modes through which a structurally diverse fiber may contribute to functional host responses.

Within this broader context, current definitions of prebiotics, including the widely cited definition proposed by the International Scientific Association for Probiotics and Prebiotics (ISAPP), provide a useful framework for interpreting interactions between dietary fibers and the gut microbiota. This definition emphasizes selective microbial utilization and associated functional effects, rather than changes in individual microbial taxa alone.(7) Framed this way, prebiotic concepts help contextualize how structurally diverse fibers engage the microbiota without implying that observed effects arise through a single microorganism or outcome.

The extent to which a fiber engages specific biological pathways may depend on intake level. At lower doses, fiber exposure may elicit detectable changes in microbial composition or activity without proportional scaling of effect size, with different taxa responding at different intakes rather than progressive amplification of the same taxa.(8–11) This dose-dependent engagement may involve distinct fermentation patterns and influence cross-feeding interactions. Thus, intake level may influence not only the magnitude, but also the nature of observed effects.

Fibers are commonly evaluated for effects on appetite and satiety; yet such outcomes represent only one dimension of fiber activity. Given the potential for structurally-complex fibers to engage multiple biological pathways, assessment of additional functional outcomes alongside appetite allows for a more comprehensive characterization of ingredient effects and their underlying biological engagement.

The present randomized, double-blind, parallel, 3-arm, placebo-controlled study was designed to examine the effects of an arabinoxylan dietary fiber over 4 to 12 weeks of supplementation at low doses (3.5g and 5g). In addition to self-reported appetite sensations, gut microbiota composition, microbial metabolite profiles, and a range of functional and metabolic outcomes were evaluated.

## 2 Materials and Methods

### 2.1 Study Design

The study was a 12-week randomized, double-blind, parallel, 3-arm, placebo-controlled clinical trial conducted at three sites in the United States (Boston, Massachusetts; Tampa Florida; Hialeah, Florida) between April 09, 2025, and October 20, 2025. The study was designed to evaluate the effects of arabinoxylan dietary fiber at low doses compared with a placebo on self-reported appetite sensations in generally healthy, overweight adults. Participants were randomly assigned in a 1:1:1 ratio to receive 1 sachet daily, of 3.5g or 5g of arabinoxylan or a placebo containing 0g of fiber.

Randomization was performed using a computer-generated algorithm. Participants were stratified by sex to ensure a balance, but not necessarily an equal number of males and females in each group. Stratification was performed manually at the sites and were electronically tracked through the electronic data capture to ensure a minimum of 45% of each sex were enrolled in each group. A subset of participants (n=90) within the randomized cohort (n=135) underwent additional study procedures for select secondary and exploratory outcomes.

The total study duration per participant was approximately 12 to 23 weeks (3-5 months), including a screening period lasting up to 75 days (to ensure regular menstruation cycles for applicable participants) and 84 days of supplementation. Study visits occurred at baseline, Week 4, and Week 12, during which questionnaires for self-reported appetite sensations, GI symptoms, and mood, as well as blood samples for blood lipids and fecal samples for microbiota were collected. In addition, at Week 4, the subset of participants had blood samples collected for glucose and insulin and individual SCFA profiles.

The study was conducted in accordance with the Declaration of Helsinki and Good Clinical Practice guidelines, and the protocol was reviewed and approved by the Sterling Institutional Review Board (Approval ID: 13170). All participants provided written consent prior to participation. The trial was registered on ClinicalTrials.gov (NCT06884449).

### 2.2 Study Population

Eligible participants were generally healthy, overweight, male and female individuals between 18 – 65 years of age (inclusive). Key inclusion criteria included a BMI between 25.0 – 29.9 kg/m^2^ (inclusive), having a habit of consuming food in the morning daily, and agree to fully consume a standardized high-carbohydrate breakfast within 15 minutes at applicable visits. Additionally, female participants either had a regular menstrual cycle, or no longer menstruated due to medication or due to being postmenopausal, surgical removal of ovaries, or medically documented ovarian failure. Key exclusion criteria included lactating, pregnant, or planning to become pregnant, participating in a weight management program or on a specific diet, or completed a weight management program within 3 months prior to screening, have current high fiber intake, experienced a change in body weight of ±4.5kg (10 lbs) over the 3 months prior to screening, have eating disorder(s), currently using lipid-lowering, anti-hypertensive, or anti-inflammatory steroid medication, or have used antibiotics, weight reducing agents, pre/probiotics, or supplements known to influence GI function within 3 months prior to screening. Participants were also required to maintain usual diet and lifestyle habits, avoid certain supplements/medications, and adhere to contraceptive requirements throughout the study. Compliance with the intervention was assessed using empty and full sachet counts and daily diaries.

Fiber intake was assessed at screening using a validated fiber intake questionnaire that included 7 questions about fruit and vegetable intake and 3 questions about foods high in fiber that rank individuals regarding their usual intake of fruits and vegetables over the past year.

Participants were recruited through the study site’s participant databases as well as print and electronic advertisement and screened for eligibility using medical history, fiber intake questionnaires, and vital signs and anthropometric measurements. A total of 183 individuals were screened, of whom 135 met eligibility criteria and were randomized to the study groups.

### 2.3 Study Intervention and Standardized Meal Consumption

To assess postprandial responses, participants came to the clinic fasted for a minimum of 10 hours, not including water, prior to the baseline, week 4, and week 12 visits for pre-breakfast assessments. Participants then consumed a standardized, high-carbohydrate breakfast in-clinic prior to blood sample collection. Post-breakfast/prandial timepoints were assessed in reference to the time of the first bite of the standardized breakfast. The breakfast provided approximately 88g of available carbohydrates, with a macronutrient composition (% total energy) of approximately 87% carbohydrate (∼90g), 10% protein (∼9g), and 3% fat (∼2g), providing approximately 390 calories and was fully consumed within 15 minutes.

At baseline, participants consumed their first dose of study intervention after all post-breakfast assessments were completed by mixing the sachet contents with 8oz of room temperature water using the provided shaker bottle. At the week 4 and 12 visits, participants consumed the dose in-clinic after all pre-breakfast assessments were completed and within 10 minutes prior to the consumption of the standardized breakfast.

Participants were instructed to follow the same dose preparation instructions for off-site dosing and to consume their dose, if possible, in the morning, with or without food.

The test products consisted of a commercially available fermentable fiber product (Arrabina®, NSC Manufacturing, UT, USA), providing 3.5g or 5g/day of wheat-derived arabinoxylan as the active ingredient. The formulation also contained small amounts of maltodextrin and flavoring agents. The maltodextrin placebo was matched for flavoring and color to match the test products.

### 2.4 Questionnaires

The primary outcome for the clinical trial was appetite sensations as measured using a visual analogue scale (VAS). The remainder of participant-reported assessments were secondary outcomes.

#### 2.4.1 Visual Analogue Scales (Appetite Sensations)

Appetite sensations were assessed using five VAS’, each with a corresponding 100 mm scale with extreme word descriptors at both ends as anchors, assessing, hunger, fullness, satiation, desire to eat food, and prospective food consumption.(12) Scale questions included “how hungry are you?”, “how full are you?”, “how satiated are you?”, “how strong is your desire to eat?”, and “how much do you think could (or would want to) to eat right now?”. Participants were instructed to draw a vertical line on the scale where they felt represented their perceived current appetite state for each question. The severity of their appetite sensation was measured using the distance (in mm) from the left-hand side of the scale to the participant’s mark on the line. Assessments were conducted at the baseline and week 4 visits and completed 15 minutes prior to breakfast and under postprandial conditions following the standardized breakfast at 5, 30, 60, 120, 180, and 240 minutes.

#### 2.4.2 GSRS (GI Symptoms)

Gastrointestinal symptoms were assessed using the Gastrointestinal Symptom Rating Scale (GSRS), a 15-item validated questionnaire comprising five subscales (abdominal pain, reflux, indigestion, diarrhea, and constipation). Each item within the subscales is rated on a 7-point Likert scale, with higher scores indicating greater symptom severity or more discomfort. Participants reported symptoms based on their experiences over the preceding 7 days. Subscale scores were calculated as the mean of the corresponding items.(13) The total score was determined by averaging all five subscale scores. Assessments were conducted at the baseline, week 4, and week 12 visits prior to the standardized breakfast.

#### 2.4.3 BRUMS-24 (Mood)

Mood states were assessed using the Brunel Mood Scale (BRUMS-24), a validated 24-item questionnaire measuring six mood domains including anger, confusion, depression, fatigue, tension, and vigor. Each domain contains 4 of the 24 questionnaire items with items rated on a 5-point scale rating ranging from 0 (not at all) to 4 (extremely). Scores for each domain were calculated by summing the score of each item within the domain to generate an overall score from 0 to 16, with higher scores indicating greater intensity of the respective mood state.(14) Participants rated how they felt “right now” (momentary recall period) at 90 minutes post-breakfast at the baseline, week 4, and week 12 visits.

### 2.5 Biological and Biochemical Assessments

Secondary outcomes included gut microbiota composition and blood lipid concentrations, while fermentation metabolite and glucose and insulin profiles were exploratory.

#### 2.5.1 Fermentation Metabolites (SCFAs)

Plasma concentrations of SCFAs including, acetic acid, propionic acid, butyric acid, pentanoic acid, hexanoic acid, and heptanoic acid were analyzed by CMBIO (Vedbæk, Denmark) and were quantified using a targeted UHPLC-MS system (Vanquish, Thermo Fisher Scientific) coupled with a high-resolution quadrupole-orbitrap mass spectrometer (Q Exactive™ HF Hybrid Quadrupole-Orbitrap, Thermo Fisher Scientific). The ionization was achieved with an electrospray ionization interface operated in negative ionization mode. Blood samples were collected at baseline and week 4. Pre-breakfast blood samples were collected within 60 minutes of the start of the standardized meal. Postprandial samples were collected at 30, 60, 120, 180, 210, and 240 minutes post-breakfast.

#### 2.5.2 Gut Microbiota Profiling

Stool samples were analyzed using 16S rRNA gene amplicon sequencing targeting the V1-V8 regions by CMBIO (Germantown, MD, USA). Fecal sample collection kits were dispensed at screening, baseline, and the week 4 visit with samples collected within 3 days of the baseline, week 4, and week 12 visits.

#### 2.5.3 Blood Biomarkers (Lipids, Glucose, Insulin)

##### 2.5.3.1 Lipids

Total cholesterol, high-density lipoprotein cholesterol (HDL-C), low-density lipoprotein cholesterol (LDL-C), and triglyceride (TG) serum concentrations were quantified using spectrophotometry by Quest Diagnostics Inc. (Boston, Massachusetts; Tampa, Florida; Hialeah, Florida). Fasted blood samples were assessed at baseline, week 4, and week 12.

##### 2.5.3.2 Glucose and Insulin

Serum fasting and postprandial glucose concentrations were quantified using spectrophotometry, while insulin was quantified using immunoassay by Quest Diagnostics Inc. (Tampa, Florida; Hialeah, Florida). Pre-breakfast blood samples were collected within 60 minutes of the start of the standardized meal. Postprandial samples were collected at 30, 60, 90, 120, 180, 210, and 240 minutes post-breakfast.

### 2.6 Safety Assessments

Safety and tolerability were evaluated throughout the study by monitoring adverse events (AEs), as well as vital signs, anthropometrics, and clinical laboratory parameters comprising of a comprehensive metabolic panel and a complete blood count with differential. AEs were collected at each study visit through open-ended questioning, spontaneous participant reporting, daily diary reporting, and investigator observation. AEs were coded using MedDRA version 28.1 and assessed for severity and relationship to the study product by the investigator.

Clinical laboratory parameters and vital signs were assessed at all visits, with vital signs measured prior to fasted blood draws. Both assessments were evaluated for clinically meaningful changes from baseline as well as abnormalities at the time of assessment. Laboratory analyses including hematology and clinical chemistry were performed by Quest Diagnostics Inc. (Boston, Massachusetts; Tampa, Florida; Hialeah, Florida) using validated methods. Concomitant medications were recorded throughout the study, and any values considered clinically significant by the investigator (e.g., elevated blood pressure readings) were followed up until resolution or stabilization.

### 2.7 Sample Size Calculation

The sample size was estimated based on the data published by Alptekin et al. 2021, in which the beta-glucan group demonstrated a Mean ± SEM AUC of postprandial prospective food consumption rating of 1063 ± 72.2 while the control had an average AUC of 1230 ± 57.2.(15) Assuming equivalent efficacy in reducing prospective food consumption rating, at 90% power, the number of participants required to complete per arm was determined to be n=27. Allowing for a 15% drop-out rate, the total number of participants required per arm was determined to be n=32. In order to account for potential differences in the efficacy of beta-glucan and low dose arabinoxylan, the sample size per arm to randomize was determined to be n=45.

### 2.8 Statistical Analysis

Statistical analyses were performed using SAS version 9.4. The primary analysis population for efficacy outcomes was the full analysis set (FAS) population, defined as all randomized participants who received at least one dose of study product and have at least one baseline efficacy assessment and one post-baseline efficacy assessment and who satisfied all inclusion/exclusion criteria upon entry (randomization). A per protocol set (PPS) population was used to corroborate the primary endpoint data. Participants were excluded from the PPS population for protocol deviations and non-compliance by the participant that could confound the evaluation of efficacy outcomes and early discontinuation of study or study product. Safety analysis was conducted in the safety set population, defined as all participants who received at least one dose of study product.

Analyses of secondary, exploratory, and post-hoc outcomes including total and individual SCFAs and glucose and insulin, were conducted in the pre-specified subset of participants (n=90) who underwent these additional assessments, as described above.

In addition, a post-hoc high-compliance population was defined for analysis based on the FAS population and included participants who completed the study and followed ≥100% dosing adherence as assessed by returned empty and full sachet counts and daily diary reporting. Analyses presented in this manuscript focus on this population to evaluate the effects of the intervention under conditions of the intended dosing regimen. Of the 125 participants in the FAS population, 100 participants were included in the high-compliance population, including 72 from the subset of 90 participants and 28 from those not included in the subset.

Where applicable, analyses were conducted within both the high-compliance population and the relevant assessment subset, resulting in outcome-specific sample sizes. Analyses conducted in the FAS are provided as secondary results for the primary and safety outcomes and supplementary material for the remaining outcomes.

Continuous variables are presented as mean ± SD, and categorical variables as frequencies and percentages. Statistical significance was assessed using two-sided tests with a significance level of α = 0.05, unless otherwise specified. For ANCOVA models, least-squares means (LSMeans) and their standard errors were estimated for each group, and between-group differences in LSMeans were tested. P-values were derived from this model.

The effects of the intervention on the primary endpoint, incremental AUC (iAUC) of VAS post-prandial appetite sensations, and exploratory endpoint, iAUC of post-prandial individual SCFAs, were analyzed using ANCOVA, with product as the fixed effect and baseline as a covariate.

For the secondary outcome of glucose and insulin, AUC was calculated and analyzed as described above.

Fasted blood lipid concentrations, question-derived scores, and microbiota composition were analyzed using ANCOVA, with product, week and interaction between product and week treated as fixed effects and subject as random effect, as well as adjustment for baseline as a covariate. Questionnaire scores were calculated according to established scoring guidelines.

Post-hoc analyses were conducted for select outcomes using the same modeling framework as the pre-specified analyses. For blood lipids, the pre-specified ANCOVA models were extended to include the additional covariate of body weight change to further explore potential influences on the observed effects. In addition, a Matsuda-derived index, triglyceride to high-density lipoprotein cholesterol ratio (TG:HDL-C), and iAUC of post-prandial composite (total) SCFA, were analyzed using the same ANCOVA approach as applied to the corresponding planned measures, with consistent model structure and covariate adjustment.

There was no special handling of missing data, and no imputation was performed for missing data unless otherwise specified.

## 3 Results

Baseline characteristics and the primary outcome are presented for the FAS and the high-compliance populations. Results for the high compliance population are presented for the remaining outcomes, unless FAS is otherwise specified.

### 3.1 Participant Flow, Baseline Characteristics, and Compliance

Between April 9 and July 24, 2025, 183 participants were screened at three sites, where an overall total of 135 participants met eligibility criteria and were randomized into the 5g arabinoxylan group (n=45), 3.5g arabinoxylan group (n=45), or placebo group (n=45). Five participants voluntarily withdrew from the study, 7 were deemed lost to follow-up, and one was withdrawn by the investigator due to non-compliance with the study product, resulting in 42, 40, and 43 participants per group, respectively, for the FAS population. Three participants were excluded from the per-protocol analysis due to early discontinuation of the study product, two participants were excluded due to out-of-window visits, and one participant was excluded due to missed doses for 3 consecutive days, resulting in 41, 38, and 40, participants in the 5g arabinoxylan, 3.5g arabinoxylan, and placebo group, respectively (**Figure 1).**

**Figure 1.**
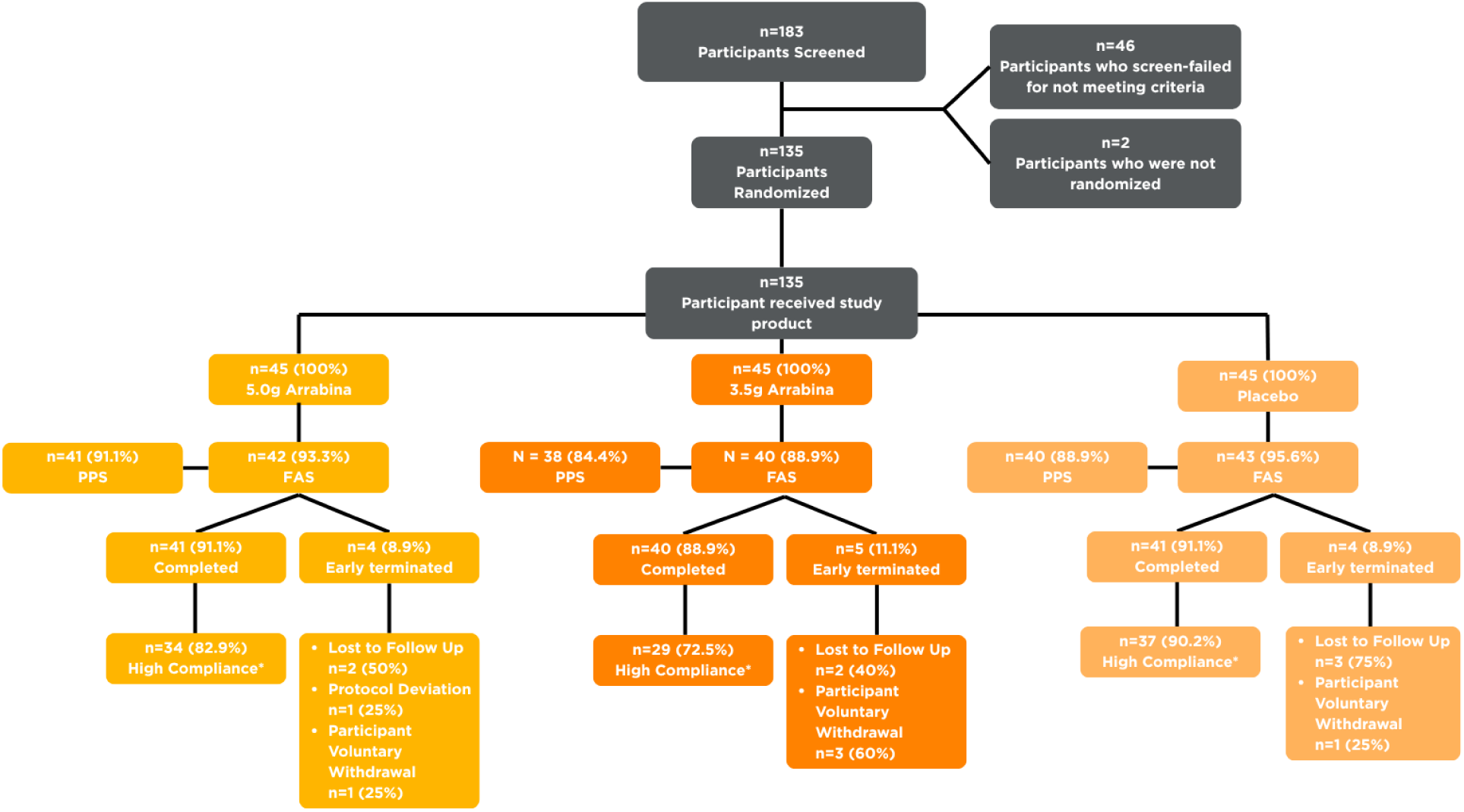
FAS, full analysis set; n, number of participants; PPS, per protocol set. CONSORT Flow diagram showing the study design and analysis populations (FAS, PPS, High-Compliance). Data are presented as n=number of participants (% of population) when percentages are specified.

Baseline demographic and anthropometric characteristics were generally comparable between groups. For the FAS population, the average age was 44.1 years with an average baseline BMI of 27.97 kg/m^2^. The participant population was diverse consisting of White (84.0%), Black or African American (13.6%), and mixed race (2.4%). Most participants were Hispanic or Latino (84.0%). There was a minimum of 45% of each sex in all groups with the overall population consisting of 48.8% females and 51.2% males (**Table 1**). The baseline demographic and anthropometric characteristics in the high-compliance population were similar to that of the FAS population (**Table 1**). Baseline characteristics were balanced between groups for both populations.

**Table 1.**
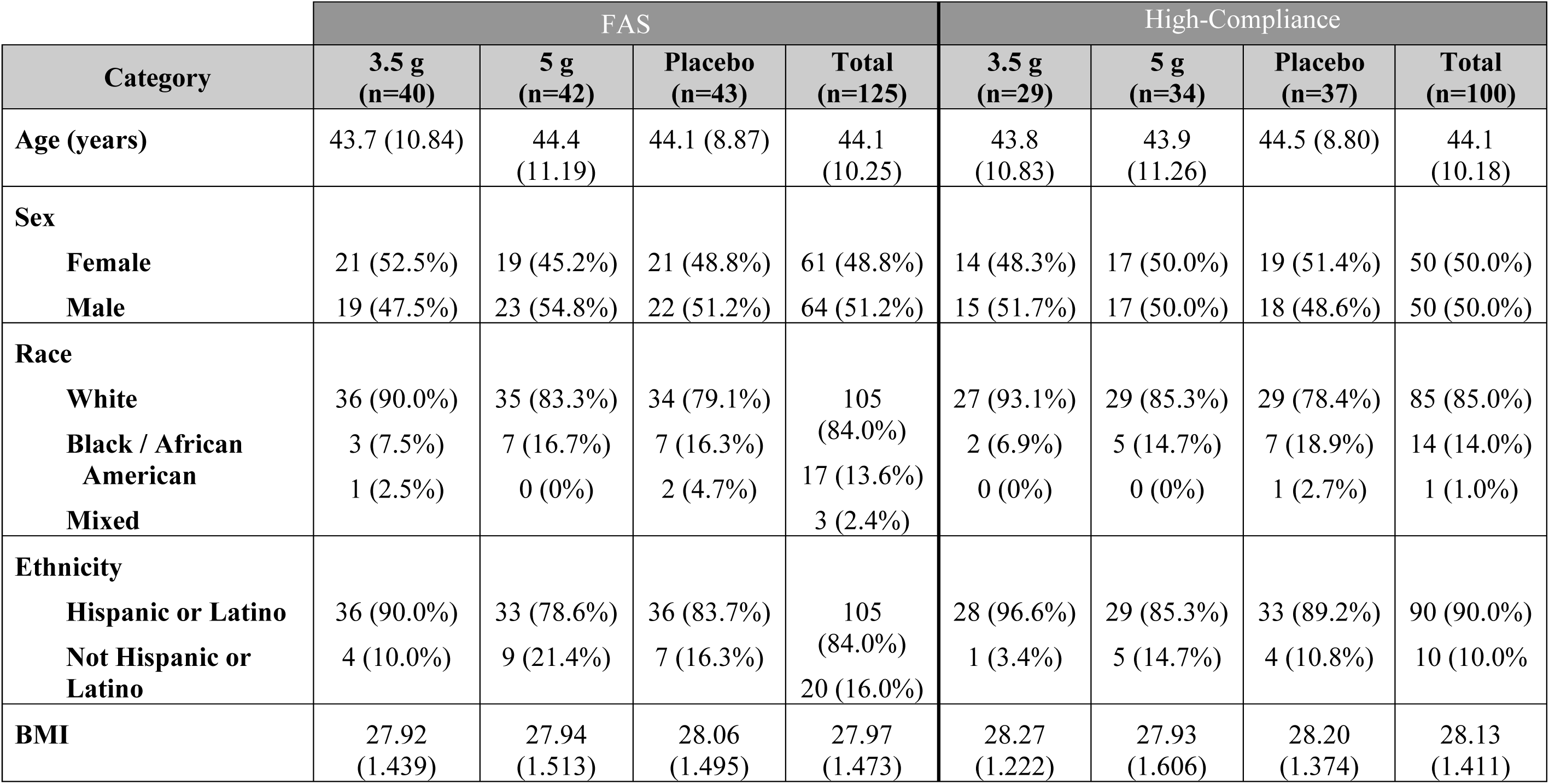
Data are presented as Mean ± SD unless percentages are specified.

The mean study product compliance across all study groups for all randomized participants for the whole study was 99.6%. Overall, for the whole study period, only one participant (5g group) had a compliance of <80% and was withdrawn from the study by the investigator before the week 4 visit.

### 3.2 Primary Outcome: Appetite Sensations

In both the high-compliance and the FAS population, there were no significant differences between groups in the change from baseline to week 4 in the iAUC of the appetite sensations measured by the VAS for either dose, including hunger, fullness, satiation, desire to eat food, and prospective food consumption (*p* > 0.05, **Table 2**). Appetite sensations at week 4 showed similar patterns in the PPS population (**Supplementary Table 1)**.

**Table 2.**
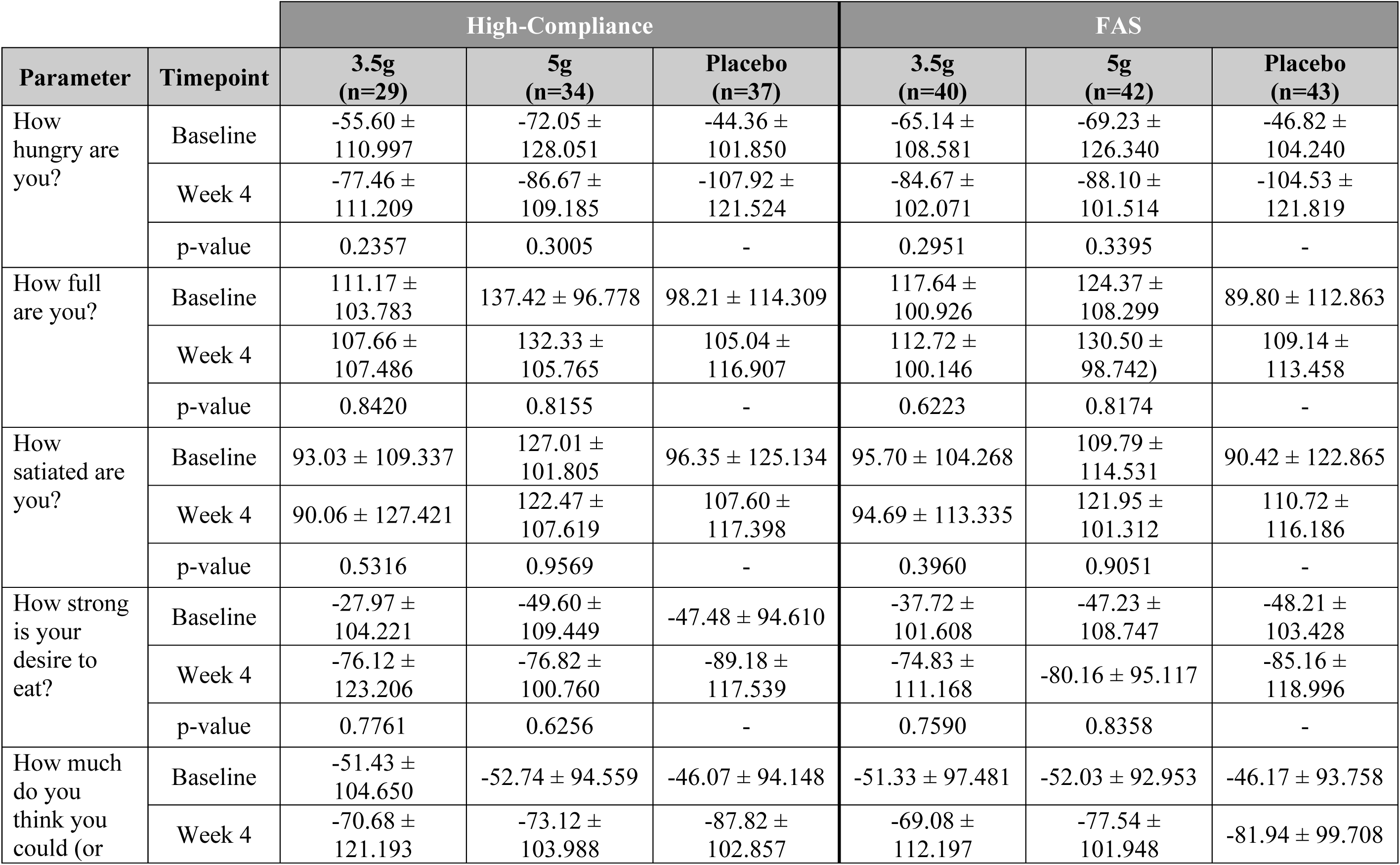
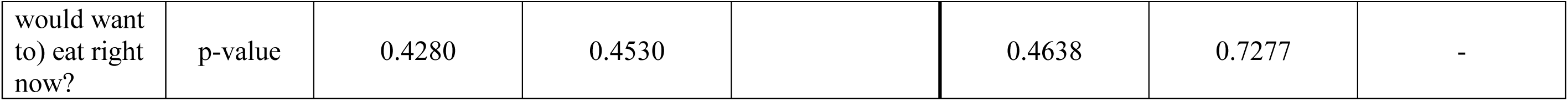
Postprandial iAUC of VAS appetite sensation rating changes from baseline to week 4. Data are presented as Mean ± SD in the high-compliance population (n=100) and FAS (n=125) for the 3.5 g and 5 g doses of arabinoxylan and placebo. Statistical significance markers are based on ANCOVA models adjusting for baseline, using least-squares means.

### 3.3 Biological Responses Following Arabinoxylan Supplementation

#### 3.3.1 Gut Microbiota Composition

Following both 4 and 12 weeks of arabinoxylan supplementation, changes were observed in select bacterial families (**Supplementary Table 2**). Relative to placebo, a significant increase in the abundance of the Lactobacillaceae family was observed at week 4 (*p* = 0.03, **Figure 2A**), while a significant increase relative to baseline in the Bacteroidaceae family was observed at week 12 (*p* = 0.02, **Figure 2B**).

**Figure 2.**
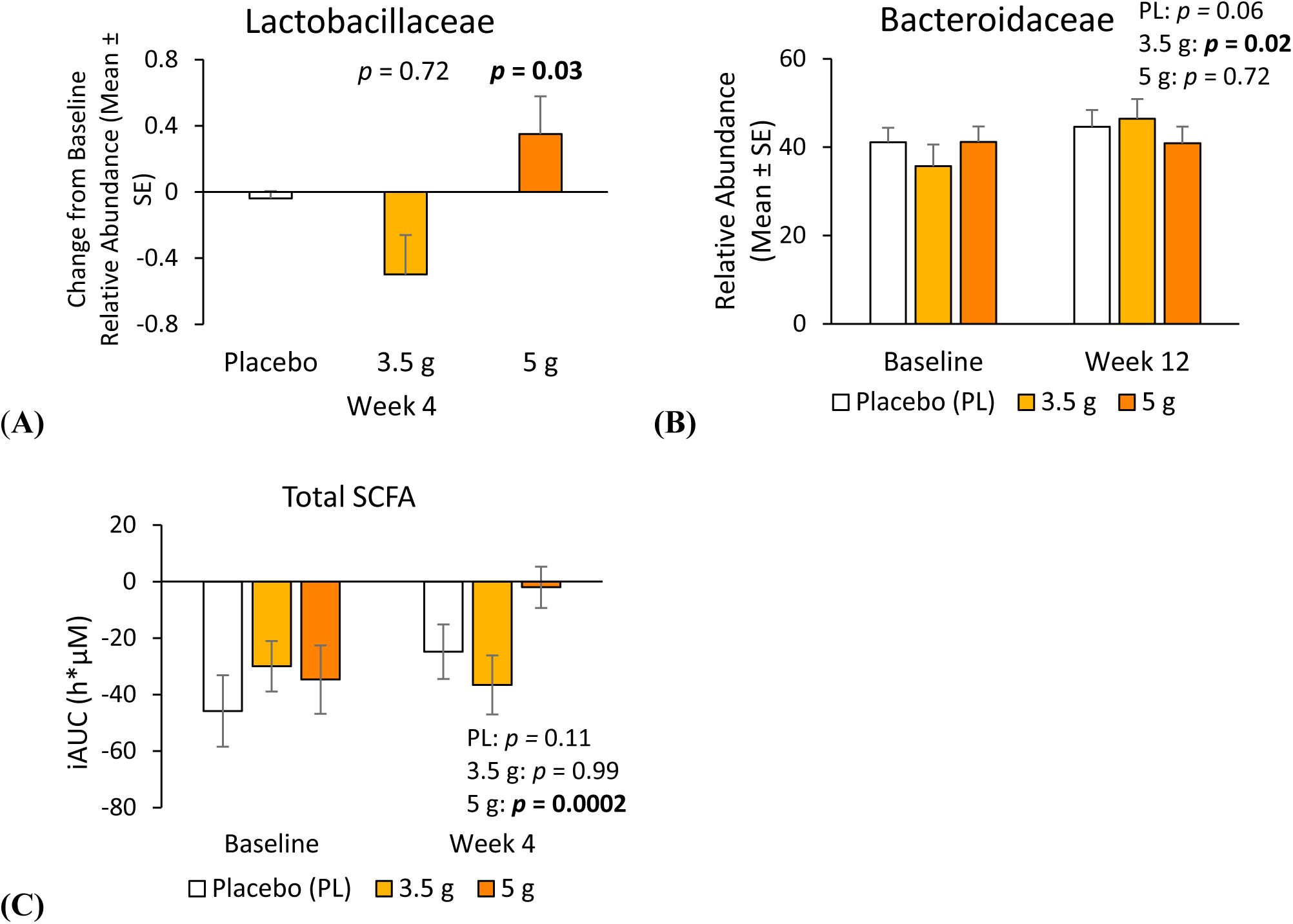
iAUC, incremental AUC; BCFA, branched-chain fatty acids; SCFA, short-chain fatty acids; SE, standard error. (A) Mean ± SE between-group, relative to placebo, change from baseline to week 4 in Lactobacillaceae abundance in the high-compliance population. (B) Mean ± SE within-group change from baseline to week 12 in Bacteroidaceae abundance in the high-compliance population. (C) Mean ± SE within-group change from baseline to week 4 in iAUC of total SCFAs in the high-compliance population.

At the genus level, significant decreases in abundance relative to placebo in *Bacteroides* were observed at week 4 (3.5g: *p =* 0.04, 5g: *p* = 0.01), while at week 12 there was an increase from baseline in *Prevotella* with the 3.5g dose (*p =* 0.02, **Supplementary Table 3**).

Data for the FAS is listed in **Supplementary Tables 2and 3**.

**Table 3.** GI symptoms assessed by GSRS total and subscale score changes from baseline to weeks 4 and 12. Data are presented as Mean ± SD in the high-compliance population (n=100) for the 3.5g and 5g doses of arabinoxylan and placebo. Bolded values are significant (*p ≤* 0.05) within-group. * Significant *p* ≤ 0.05, ** Significant *p* ≤ 0.01, *** Significant *p* < 0.001 compared to placebo. Statistical significance markers are based on ANCOVA models adjusting for baseline, using least-squares means.

| Parameter | Timepoint | 3.5g<br>(n=29) | 5g<br>(n=34) | Placebo<br>(n=37) |
| --- | --- | --- | --- | --- |
| **Subscale** |  |  |  |  |
| Abdominal Pain | Baseline | 1.2874 ± 0.5615 | 1.1667 ± 0.2752 | 1.1441 ± 0.2782 |
| | Week 4 | 1.1609 ± 0.3844 | 1.2353 ± 0.4752 | 1.2703 ± 0.4833 |
| | Week 12 | **1.0690 ± 0.1864** | 1.2255 ± 0.4398 | 1.1982 ± 0.4191 |
| Constipation | Baseline | 1.3103 ± 0.7011 | 1.2549 ± 0.5921 | 1.3063 ± 0.5740 |
| | Week 4 | **1.1034 ± 0.2536** | 1.2451 ± 0.5210 | 1.2162 ± 0.4793 |
| | Week 12 | 1.1494 ± 0.4763 | 1.1765 ± 0.3599 | 1.2973 ± 0.7769 |
| Diarrhea | Baseline | 1.0690 ± 0.2250 | 1.0294 ± 0.0960 | 1.1081 ± 0.3690 |
| | Week 4 | 1.1034 ± 0.3225* | 1.1667 ± 0.4732 | **1.3153 ± 0.6332** |
| | Week 12 | 1.0230 ± 0.0860 | 1.0980 ± 0.3234 | 1.1081 ± 0.2945 |
| Indigestion | Baseline | 1.276 ± 0.6955 | 1.228 ± 0.3712 | 1.405 ± 0.5476 |
| | Week 4 | 1.293 ± 0.6376 | 1.228 ± 0.4742 | 1.399 ± 0.5787 |
| | Week 12 | 1.164 ± 0.3490 | 1.228 ± 0.4191 | 1.324 ± 0.5889 |
| Reflux | Baseline | 1.16 ± 0.502 | 1.03 ± 0.119 | 1.12 ± 0.447 |
| | Week 4 | 1.07 ± 0.221 | 1.01 ± 0.086* | 1.18 ± 0.338 |
| | Week 12 | 1.05 ± 0.205 | 1.16 ± 0.363 | 1.14 ± 0.326 |
| **Total** |  |  |  |  |
| Total | Baseline | 1.2195 ± 0.4761 | 1.1417 ± 0.1911 | 1.2171 ± 0.2990 |
| | Week 4 | 1.1460 ± 0.2484* | 1.1779 ± 0.2846 | 1.2752 ± 0.3342 |
| | Week 12 | **1.0914 ± 0.2035** | 1.1779 ± 0.2413 | 1.2126 ± 0.3718 |

#### 3.3.2 Fermentation Products (SCFAs)

##### 3.3.2.1 Planned Analysis: Individual SCFA

Significant increases from baseline were observed in the iAUC of acetic acid (*p* = 0.002) and pentanoic acid (*p* = 0.04, **Supplementary Table 4**) following 4 weeks of supplementation at the 5g dose, although between-group comparisons relative to placebo were not significant.

In the FAS, SCFA measures showed similar directional changes to those observed in the high-compliance population (**Supplementary Table 4**).

##### 3.3.2.2 Post-hoc Analysis: Total SCFAs

Changes in overall bacterial metabolite concentrations within the 5g group were observed over the 4-week intervention period where the iAUC of total SCFAs significantly increased from baseline (*p* = 0.0002, **Figure 2C**) although comparison to placebo did not reach significance (**Supplementary Table 4**).

Comparisons to baseline and placebo for total SCFA in the FAS did not reach statistical significance.

### 3.4 Functional Outcomes Following Arabinoxylan Supplementation

#### 3.4.1 GI Comfort (GSRS)

Changes were observed in multiple GSRS subscales over the intervention period (**Table 3**). At week 4, a significant reduction in GSRS total score was observed in the 3.5g intervention arm relative to placebo (*p* = 0.05, **Figure 3A**), as well as a within-group reduction at week 12 (*p* = 0.03) that did not reach significance relative to placebo. Significant within-group reductions from baseline were also observed for constipation at week 4 (*p =* 0.03) and abdominal pain at week 12 (*p* = 0.05), although between-group differences for these scales did not reach statistical significance. In addition, a significant reduction in reflux symptoms relative to placebo was observed at week 4 in the 5g intervention arm (*p* = 0.02, **Figure 3B**).

**Figure 3.**
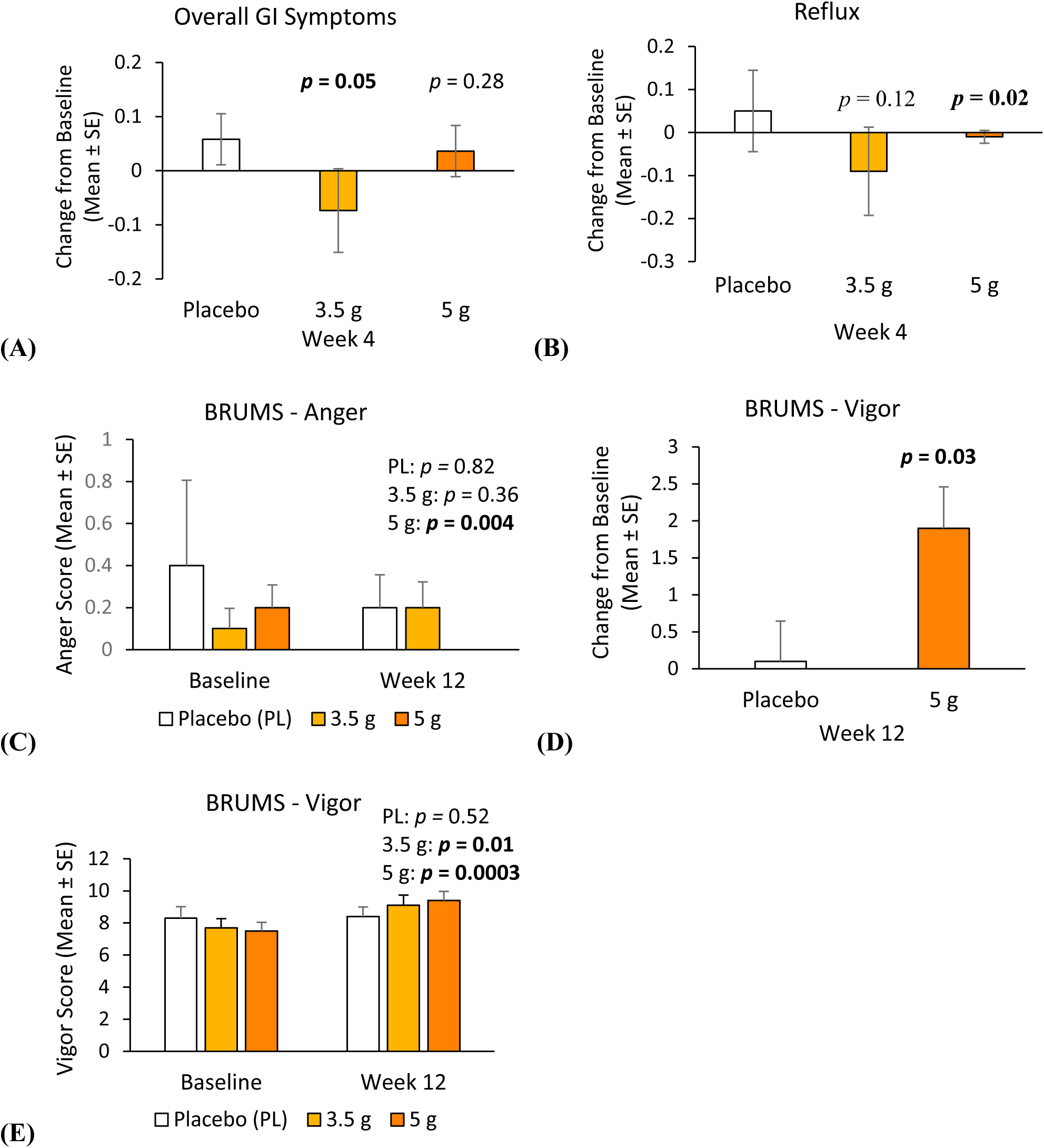
BRUMS, brunel mood scale; GI, gastrointestinal; GSRS, gastrointestinal symptoms rating scale; SE, standard error. (A) Mean ± SE between-group, relative to placebo, change from baseline to week 12 in total GSRS score in the high-compliance population. (B) Mean ± SE between-group, relative to placebo, change from baseline to week 4 in GSRS reflux score in the high-compliance population. (C) Mean ± SE within-group, change from baseline to week 12 in BRUMS anger score in the high-compliance population. (D) Mean ± SE between-group, relative to placebo, change from baseline to week 12 in BRUMS vigor score of 5g group in the high-compliance population. (E) Mean ± SE within-group change from baseline to week 12 in BRUMS vigor score in the high-compliance population.

In the FAS population, GSRS total and scale scores did not differ significantly from placebo and did not show consistent within-group changes over the intervention period (**Supplementary Table 5**).

#### 3.4.2 Mood (BRUMS-24)

Changes were observed in select mood domains over the intervention period. A significant within-group decrease in anger was observed in the 5g arm at week 4 (*p* = 0.05) and week 12 (*p =* 0.004, **Figure 3C**); however, the between-group comparison relative to placebo at week 4 was not statistically significant, while the week 12 comparison was (*p* = 0.05). Additionally, a significant increase in the vigor domain (*p* = 0.03, **Figure 3D**) relative to placebo, was also observed at week 12 (**Table 4**).

**Table 4.** Mood assessed by BRUMS-24 domain score changes from baseline to weeks 4 and 12. Data are presented as Mean ± SD in the high-compliance population (n=100) for the 3.5g and 5g doses of arabinoxylan and placebo. Bolded values are significant (*p ≤* 0.05) within-group. * Significant *p* ≤ 0.05, ** Significant *p* ≤ 0.01, *** Significant *p* < 0.001 compared to placebo. Statistical significance markers are based on ANCOVA models adjusting for baseline, using least-squares means.

| Parameter | Timepoint | 3.5g<br>(n=29) | 5g<br>(n=34) | Placebo<br>(n=37) |
| --- | --- | --- | --- | --- |
| Anger | Baseline | 0.1 ± 0.52 | 0.2 ± 0.63 | 0.4 ± 2.47 |
|  | Week 4 | **0.0 ± 0.00** | **0.1 ± 0.38** | **0.0 ± 0.00** |
| | Week 12 | 0.2 ± 0.66 | **0.0 ± 0.00\*** | 0.2 ± 0.95 |
| Confusion | Baseline | 0.1 ± 0.41 | 0.2 ± 0.46 | 0.3 ± 0.82 |
| | Week 4 | 0.1 ± 0.31 | 0.2 ± 0.72 | 0.1 ± 0.39 |
| | Week 12 | 0.2 ± 0.68 | 0.1 ± 0.69 | 0.4 ± 1.28 |
| Depression | Baseline | 0.2 ± 0.79 | 0.2 ± 0.63 | 0.4 ± 1.21 |
| | Week 4 | 0.4 ± 1.08 | 0.1 ± 0.70 | 0.3 ± 1.18 |
| | Week 12 | 0.3 ± 1.49 | 0.1 ± 0.69 | 0.3 ± 0.87 |
| Fatigue | Baseline | 1.7 ± 2.37 | 1.4 ± 2.09 | 1.9 ± 2.45 |
| | Week 4 | 1.9 ± 3.06 | 1.8 ± 2.68 | 2.2 ± 2.93 |
| | Week 12 | 1.1 ± 1.39 | 1.2 ± 1.77 | 2.1 ± 2.40 |
| Tension | Baseline | 0.6 ± 1.06 | 0.5 ± 1.40 | 0.6 ± 1.09 |
| | Week 4 | 0.6 ± 1.02 | 0.4 ± 1.10 | 0.5 ± 0.93 |
| | Week 12 | 0.4 ± 1.35 | 0.5 ± 1.46 | 0.4 ± 0.90 |
| Vigor | Baseline | 7.7 ± 3.08 | 7.5 ± 3.16 | 8.3 ± 4.34 |
| | Week 4 | 8.0 ± 3.23 | 8.0 ± 3.39 | 8.3 ± 3.96 |
| | Week 12 | **9.1 ± 3.45** | **9.4 ± 3.32\*** | 8.4 ± 3.64 |

In the 3.5g intervention arm, a significant within-group decrease in the anger domain (*p =* 0.007) was observed at week 4, and a significant within-group increase in the vigor domain (*p* = 0.01; **Figure 3E**) was observed at week 12; neither finding reached statistical significance compared with placebo. No consistent changes were observed in the remaining domains at either timepoint, including confusion, depression, fatigue, and tension.

BRUMS mood responses for the FAS followed patterns consistent with those observed in the high-compliance population (**Supplementary Table 6**).

#### 3.4.3 Blood Lipids

#### 3.4.3.1 Planned Analysis: Lipid Concentrations

Significant reductions in total cholesterol concentration from baseline were observed following supplementation with 3.5g at week 12 (*p =* 0.01, **Figure 4A**, **Table 5**). Compared to placebo, significant reductions in triglyceride concentrations (*p =* 0.04, **Figure 4B**) and LDL-C (*p* = 0.05, **Figure 4C**) were observed in the 3.5g group. Significant reductions in HDL-C with both doses relative to placebo were observed at week 4 (3.5g: *p* = 0.02, 5g: *p* = 0.007); however, this effect was not maintained at week 12 (*p* > 0.05, **Figure 4D)**.

**Figure 4.**
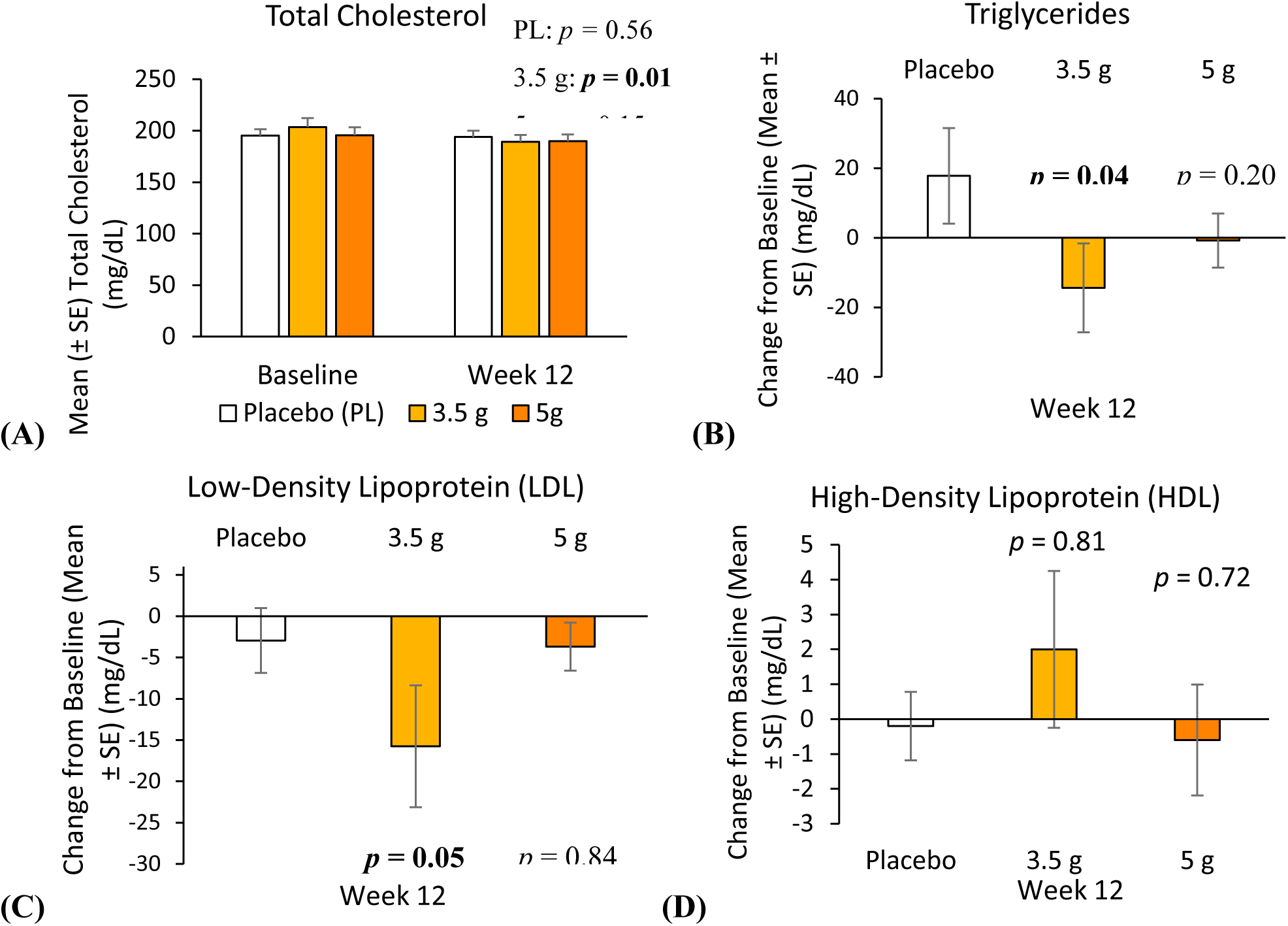
SE, standard error. (A) Mean ± SE within-group change from baseline to week 12 in total cholesterol in the high-compliance population. (B) Mean ± SE between-group, relative to placebo, change from baseline to week 12 in triglycerides in the high-compliance population. (C) Mean ± SE between-group, relative to placebo, change from baseline to week 12 in LDL-C in the high-compliance population. (D) Mean ± SE between-group, relative to placebo, change from baseline to week 12 in HDL-C in the high-compliance population.

**Table 5.**
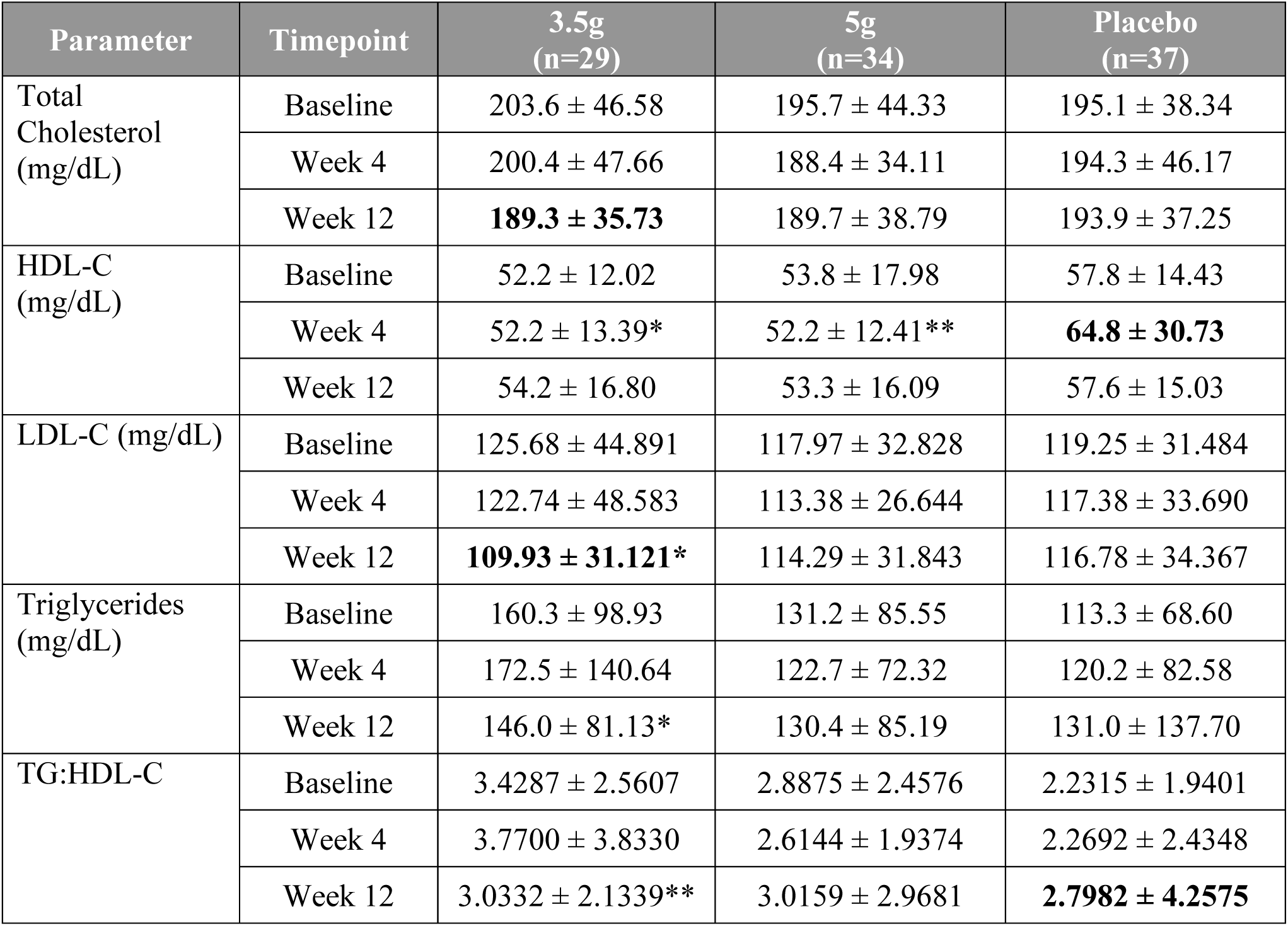
HDL-C, high-density lipoprotein cholesterol; LDL-C, low-density lipoprotein cholesterol; TG, triglycerides. Blood lipid concentration and TG:HDL-C ratio changes from baseline to weeks 4 and 12. Data are presented as Mean ± SD in the high-compliance population (n=100) for the 3.5g and 5g doses of arabinoxylan and placebo. Bolded values are significant (*p ≤* 0.05) within-group. * Significant *p* ≤ 0.05, ** Significant *p* ≤ 0.01, *** Significant *p* < 0.001 compared to placebo. Statistical significance markers are based on ANCOVA models adjusting for baseline, using least-squares means.

Lipid responses in the FAS population demonstrated partial overlap with patterns observed in the high-compliance population (**Supplementary Table 7**).

##### 3.4.3.2 Post-hoc Analysis: TG:HDL-C Ratio

In a post-hoc analysis, a significant reduction in the TG:HDL-C ratio compared to placebo was observed at week 12 in both the high-compliance (*p* = 0.01, **Table 5**) and FAS population (*p* = 0.03, **Supplementary Table 7**) receiving the 3.5g intervention.

##### 3.4.3.3 Post-hoc Analysis: Weight-Adjusted Sensitivity Analysis

In a post-hoc sensitivity analysis adjusting for change in body weight, additional significant within-group reductions in total cholesterol were observed in the 5g group at both week 4 (*p* = 0.02) and week 12 (*p* = 0.05, **Supplementary Table 8**), which were not evident in previous analyses. In contrast, total cholesterol changes at the 3.5g dose at week 12 and HDL-C changes relative to placebo across doses were consistent with findings from prior analyses. Similarly, reductions in LDL-C and triglycerides at week 12 in the 3.5g intervention arm aligned with results from the high-compliance analysis. Adjusting for body weight did not yield any new results in the FAS population (**Supplementary Table 8**).

### 3.5 Metabolic Outcomes Following Arabinoxylan Supplementation

#### 3.5.1 Planned Analysis: Glucose and Insulin

The AUC of both glucose (*p* = 0.01) and insulin (*p* = 0.03) significantly increased from baseline in the 3.5g intervention arm at 4 weeks; however, only the between-group comparison relative to placebo for glucose reached statistical significance (*p* = 0.02, **Supplementary Table 9**). No statistically significant changes in either AUC were observed in the 5g intervention arm.

In the full analysis set at week 4, a significant within-group increase in insulin AUC (*p* = 0.05) was observed in the 3.5g arm, whereas no significant changes from baseline were detected for glucose or for either parameter in the alternate intervention compared with placebo (**Supplementary Table 9**).

#### 3.5.2 Post-hoc Analysis: Matsuda-Derived Index

Derived measures of overall glucose and insulin exposure, estimated using a Matsuda-inspired index, did not demonstrate statistically significant changes in the high-compliance population (**Supplementary Table 9**).

For the FAS, a significant increase from baseline was observed at week 4 in the 5g intervention arm (*p* = 0.01), while no statistically significant differences relative to placebo were detected (**Supplementary Table 9**).

### 3.6 Safety

Overall, arabinoxylan supplementation in the current formulation at low doses demonstrated good safety and tolerability profiles with no SAEs or deaths or dropouts due to a treatment-emergent adverse event (TEAE). TEAEs were mainly mild in severity. Most TEAEs that were possibly related to the study products occurred in the placebo group (**Table 6**). At the end of the study, all TEAEs were resolved.

**Table 6.** %, percentage of participants having experienced an AE; AE, adverse event; E, number of AEs; n, number of participants; S, number of participants having experienced an AE. Data is presented as Number of participants (Percentage of participants having experienced an AE) number of AEs. Safety analysis was conducted in the Safety Set population.

|  | 3.5g<br>(n=45) | 5g<br>(n=45) | Placebo<br>(n=45) | Total<br>(n=135) |
| --- | --- | --- | --- | --- |
|  | S (%) E |  |  |  |
| Total AE Reported |  |  |  |  |
|  | 6 (13.3%) 10 | 6 (13.3%) 15 | 5 (11.1%) 18 | 17 (12.6%) 43 |
| Relation |  |  |  |  |
| Related | 0 (0%) 0 | 0 (0%) 0 | 0 (0%) 0 | 0 (0%) 0 |
| Possibly related | 1 (2.2%) 2 | 3 (6.7%) 6 | 3 (6.7%) 10 | 7 (5.2%) 18 |
| Unlikely related | 0 (0%) 0 | 1 (2.2%) 1 | 2 (4.4%) 4 | 3 (2.2%) 5 |
| Not related | 5 (11.1%) 8 | 2 (4.4%) 8 | 0 (0%) 4 | 7 (5.2%) 20 |
| Severity |  |  |  |  |
| Severe | 0 (0%) 0 | 0 (0%) 0 | 0 (0%) 0 | 0 (0%) 0 |
| Moderate | 0 (0%) 0 | 2 (4.4%) 2 | 0 (0%) 0 | 2 (1.5%) 2 |
| Mild | 6 (13.3%) 10 | 4 (8.9%) 13 | 5 (11.1%) 18 | 15 (11.1%) 41 |
| Serious |  |  |  |  |
|  | 0 (0%) 0 | 0 (0%) 0 | 0 (0%) 0 | 0 (0%) 0 |
| Leading to Discontinuation |  |  |  |  |
|  | 0 (0%) 0 | 0 (0%) 0 | 0 (0%) 0 | 0 (0%) 0 |

## 4 Discussion

The present study evaluated the biological and functional responses to low doses of arabinoxylan across GI, metabolic, behavioral, and microbial domains in healthy, overweight adults under free-living conditions. Rather than demonstrating a singular or uniform mechanism of action, the findings characterize the ingredient as a biologically active and versatile fiber capable of engaging multiple systems, with domain-specific patterns of response that were influenced by exposure and inter-individual variability. Collectively, the data supports selective biological activity consistent with prebiotic-like behavior while underscoring the complexity and context-dependence of human dietary responses.

Intake of low-dose arabinoxylan, daily for 4 to 12 weeks, was associated with within-group changes in select microbial fermentation products, including SCFAs, as well as significant shifts in specific microbial taxa, in the high-compliance population (compliance greater than or equal to 100%), indicating engagement of the gut microbiota under conditions of consistent, repeated exposure. SCFA responses were assessed using iAUC, capturing the net within-day, post-prandial change in circulating concentrations and reflecting the dynamic, time-integrated nature of microbial fermentation and metabolite availability. The observed changes were taxon- and metabolite-specific rather than global in nature, suggesting selective microbial interactions rather than broad restructuring of the microbial community.

Specifically, increases in Lactobacillaceae and Bacteroidaceae families, as well as *Prevotella* at the genus level, alongside greater cumulative SCFA responses (iAUC), including acetate and pentanoate, were observed. Members of these microbial families are commonly associated with the fermentation of complex carbohydrates and the production of SCFAs, particularly acetate.(16,17) The parallel changes in microbial taxa and integrated SCFA responses over the sampling period suggest coordinated shifts in microbial metabolic activity, consistent with enhanced fermentation of the dietary substrate following repeated exposure. Importantly, the use of iAUC indicates that these effects reflect an increased overall post-prandial SCFA response rather than isolated changes at individual timepoints, supporting a functional engagement of the gut microbiota.

Additionally, a decrease in *Bacteroides* abundance relative to placebo at week 4 was observed. Responses of *Bacteroides* to dietary interventions have been inconsistently reported in the literature and appear highly context-dependent, influenced by baseline microbiota composition, fiber structure, and host factors. Several controlled fiber studies have reported both increases and decreases in *Bacteroides* across interventions, reinforcing the view that genus-level changes should be interpreted cautiously and not assumed to reflect uniform functional consequences.(18–20) Accordingly, these *Bacteroides* findings are best understood as indicators of selective microbial changes rather than direct drivers of host outcomes.

The absence of widespread or uniform microbial shifts aligns with evidence from human fiber studies demonstrating that dietary fibers often elicit targeted shifts in microbial activity and metabolite production without consistent or large-scale changes in overall community composition. Meta-analyses and re-analyses of fiber interventions indicate that microbial responses are dominated by inter-individual variability, with fiber intake explaining only a small proportion of overall compositional variation, despite reproducible changes in select taxa and metabolic outputs.(18,21)

Beyond microbial engagement, intake of the fiber was associated with domain-specific functional outcomes. Improvements in GI comfort, reflected by changes in select GSRS scores, were most apparent under conditions of sustained intake, consistent with the expected physiological role of dietary fibers in the gut environment. These responses were not accompanied by changes in appetite sensations, suggesting that GI effects were not driven by alterations in hunger or satiety perception.

Behavioral outcomes also demonstrated selective patterns. Changes were observed in BRUMS anger and vigour domains at week 12, while other mood dimensions remained largely unchanged. The concurrent changes observed in these domains suggest an effect on mood regulation and may reflect coordinated modulation of affective processes over time. Notably, these changes occurred independently of appetite effects, supporting the interpretation that observed behavioral responses were not secondary to energy intake regulation.

Lipid outcomes further illustrate the ingredient’s capacity for system-specific effects, particularly under conditions of continued daily dosing. Reductions in LDL-C and triglycerides relative to placebo were observed in the high-compliance population at the 3.5g intake level, with more variable responses in the FAS population. The test product’s effects on lipid outcomes in the high-compliance population, while examined post-hoc, provides insight into lipid responses under intended, daily consumption rather than serving as a confirmatory efficacy analysis.

Changes in total cholesterol relative to baseline were observed as well, as expected given the contribution of these fractions to total cholesterol, and did not provide an independent signal beyond the primary lipid components. Sensitivity analyses adjusting for body weight did not materially alter the interpretation of lipid outcomes, supporting the robustness of the observed lipid pattern.

In addition to individual lipid parameters, the TG:HDL-C ratio demonstrated a favorable pattern with consistent daily dosing. As a composite marker reflecting the balance between atherogenic and protective lipid fractions, this ratio has been associated with insulin resistance and cardiometabolic risk.(22) In the present study, changes in the TG:HDL-C ratio were directionally consistent with the reductions observed in triglycerides, supporting the overall lipid profile shift rather than representing an independent signal. While these findings should be interpreted cautiously given the exploratory nature of this endpoint, they provide additional context for the coordinated lipid responses observed and further support the potential cardiometabolic relevance of the intervention under sustained intake conditions.

The lipid responses occurred alongside evidence of microbial fermentation and selective microbiota engagement, which provides biological plausibility within the framework of established dietary fiber-lipid interactions. Evidence from randomized controlled trials and meta-analyses demonstrate that fermentable and soluble dietary fibers are associated with consistent improvements in circulating lipids through integrated effects on cholesterol metabolism and bile acid handling.(23,24) In this context, microbiota changes are thought to reflect shifts in substrate utilization and fermentation activity rather than acting as direct drivers of lipid outcomes. Fermentation metabolites such as SCFAs have been implicated in cholesterol regulatory pathways in both mechanistic and human supplementation studies, providing further contextual support for the observed lipid pattern.(25,26)

Collectively, the findings indicate that LDL-cholesterol and triglycerides are responsive cardiometabolic endpoints under sustained intake conditions, with total cholesterol changes providing supportive context. These results support further investigation of the ingredient’s cardiometabolic potential in studies specifically designed and powered to evaluate lipid outcomes.

Glycemic measures, including glucose and insulin AUC, exhibited substantial variability and did not demonstrate consistent between-group difference relative to placebo. However, exploratory analysis using a Matsuda-derived index suggested altered overall glucose-insulin exposure in the FAS population. This index was included to provide an integrated assessment of glucose-insulin dynamics that may capture broader metabolic patterns not evident in individual measures. However, given the derived and post-hoc nature of this index, and the absence of corresponding signals in primary glycemic endpoints, this finding should be interpreted as hypothesis-generating.

The observed combination of selective microbial changes accompanied by host-relevant functional responses aligns with key elements of the ISAPP definition of a prebiotic: a substrate that is selectively utilized by host microorganisms conferring a health benefit.(7) In the present study, evidence of microbial fermentation and taxon-specific shifts is consistent with selective utilization, as reflected by the parallel increases in Lactobacillaceae and Bacteroidaceae alongside elevations in cumulative composite and select SCFAs. These microbial and metabolic changes occurred in parallel with domain-specific host responses, including alterations in lipid parameters and other functional outcomes, supporting the presence of a host-relevant, microbiota-mediated effect. Importantly, these findings do not suggest a singular microbiota-driven mechanism, but rather support a broader, modern interpretation in which prebiotic activity may manifest through multiple, interacting biological pathways. Given the variability observed and the influence of placebo responsiveness, replication would strengthen confidence in these interpretations.

SCFAs, produced via fermentation of dietary fibers by certain taxa, have been connected to host metabolic regulation through several complementary pathways. Following absorption, SCFAs may enter portal circulation and reach the liver where they may influence lipid homeostasis and cholesterol metabolism. Acetate, in particular, has been associated with modulation of hepatic lipid metabolism, including reduced lipogenesis through regulatory pathways involving PPARα and AMP-activated protein kinase (AMPK), as well as increased fatty acid oxidation.(27,28) These effects provide a biologically plausible pathway through which fermentation-derived metabolites may contribute to the reductions in circulating triglycerides and LDL-cholesterol observed in the present study.

In addition to hepatic effects, SCFAs can act as signaling molecules via G protein–coupled receptors (e.g., FFAR2/FFAR3), where they may influence adipose tissue metabolism. Activation of these pathways has been associated with inhibition of lipolysis and reduced release of circulating free fatty acids, thereby limiting substrate availability for hepatic triglyceride synthesis.(27,29) Through this mechanism, SCFA signaling may contribute to systemic regulation of energy balance and lipid handling, providing an additional pathway linking microbial fermentation to circulating lipid outcomes.

At a broader system level, lipid metabolism may also be influenced through interactions within the gut-liver axis involving bile acid signaling. Gut microbiota, such as the Lactobacillaceae family, have been implicated in bile acid transformation and regulation, which can influence cholesterol homeostasis and lipid transport.(30) These bile acid–mediated pathways, alongside SCFA-driven metabolic effects, provide a complementary framework through which microbial activity may contribute to the lipid-related outcomes observed in the present study. However, these relationships were not directly assessed and should be interpreted as coordinated system-level responses rather than evidence of a defined mechanistic pathway.

Additionally, the changes in SCFAs, assessed via iAUC, indicate that the test product modulated the acute, postprandial profile of microbial fermentation products. This approach captures the cumulative response relative to baseline, providing insight into net SCFA production and/or absorption dynamics rather than single timepoint fluctuations. The selective nature of the SCFA response suggests that the test product may influence specific metabolic pathways or substrate utilization patterns within the gut microbiota, rather than inducing a generalized increase in fermentation activity. These findings support a targeted effect of the test product on microbial metabolic function.

Furthermore, while the study was not designed to investigate gut-brain mechanisms and mechanistic conclusions cannot be drawn from the present data, these findings are directionally consistent with growing evidence that dietary fiber intake and microbial activity may influence affective domains via microbiota-gut-brain communication pathways. Current human evidence supports modest and context-dependent associations between fiber intake and mood-related outcomes.(31,32)

Focusing on the high-compliance population, examined post-hoc, provided insight into biological responses under conditions of once daily intake, as per the intended dosing regimen. Findings in this subgroup did not contradict those observed in the full analysis set but reflected more coherent expression of similar patterns, underscoring the influence of exposure consistency in dietary fiber research.

At the same time, within-group changes were observed in the placebo arm that paralleled the direction of responses hypothesized for the test product, indicating a measurable placebo contribution. Dietary fiber is commonly associated with perceived benefits for digestive comfort and appetite regulation, which may influence participant expectations when entering an intervention study. Expectancy effects are known to disproportionately affect questionnaire-based outcomes, which are inherently sensitive to individual interpretation(33), particularly in populations with elevated health awareness or motivation for change.(34) In this context, pre-existing beliefs regarding the effects of fiber may have contributed to improvements observed in the placebo group across select subjective endpoints. The coexistence of placebo effects alongside selective ingredient-associated changes highlights the complexity of multisystem dietary interventions.

Strengths of the study include comprehensive assessment across biological and functional domains, additional analysis under sustained-exposure conditions (high compliance), and transparent reporting of exploratory and sensitivity analyses. Limitations include heterogeneity of responses, the inherent sensitivity of microbiota endpoints to contextual factors, and the analysis of fermentation metabolites and metabolic biomarker analysis only in a subset of participants, which may limit generalizability and statistical power for these outcomes. These factors highlight the importance of cautious interpretation and the need for further research.

In conclusion, the low doses of arabinoxylan evaluated in this study demonstrated biologically selective, domain-specific activity across microbial, GI symptom, metabolic, and behavioral outcomes. These effects were most coherently expressed under conditions of consistent daily intake and occurred independently of changes in appetite sensation. Rather than supporting a singular mechanism, the findings highlight the ingredient’s biological versatility and contextual responsiveness and suggest prebiotic potential through alignment with the ISAPP definition, supporting further investigation to better characterize the mechanistic pathways underlying these observations.

## 6 Author Contributions

**ACA:** Writing - review & editing, Writing - original draft. **JW:** Writing - review & editing, Formal analysis. **S-AG:** Writing - review & editing, Methodology. **KAF:** Writing - review & editing, Visualization. **AA:** Writing - review & editing, Project administration, Resources, Supervision. **JB:** Writing - review & editing, Conceptualization, Methodology. **SR:** Writing - review & editing, Methodology, Project administration, Supervision. **HSA:** Writing - review & editing. **AR:** Conceptualization, Methodology, Writing - review & editing

## 8 Acknowledgments

The authors would like to thank Bruce Hamaker, Kristin Verbeke, and Edward Deehan of Comet’s scientific advisory board, for their contribution to the conceptualization of the study design, the Principal Investigators: Dr. Allen Kuhn (Vantage Clinical Trials, Tampa, FL, United States), Dr. Jose Luis Vega (Indago Research, Hialeah, FL, United States), and Dr. Antonio Mendes (Boston Clinical Trials, Boston, MA, United States) and their study staff for their study conduct, as well as the study participants for their involvement.

## 10 Data Availability Statement

The datasets analyzed for this study are available upon request.

