## Supplemental Tables 1 to 9 for "Supplementation with Arabinoxylan Dietary Fiber at Low Doses Produces Behavioral, Metabolic, and Gut Microbial Changes in Healthy, Overweight Adults: A Randomized Placebo-Controlled Trial"

**Supplementary Table 1** Postprandial iAUC of VAS appetite sensation rating changes from baseline to week 4. Data are presented as Mean ± SD in the PPS (n=119) for the 3.5g and 5g doses of arabinoxylan and placebo. Statistical significance markers are based on ANCOVA models adjusting for baseline, using least-squares means.

| | | PPS | | |
|---|---|---|---|---|
| **Parameter** | **Timepoint and p-value** | **3.5g (n=38)** | **5g (n=41)** | **Placebo (n=40)** |
| How hungry are you? | Baseline | -63.43 ± 110.771 | -76.18 ± 119.496 | -45.51 ± 104.879 |
| | Week 4 | -83.17 ± 104.499 | -88.88 ± 102.649 | -110.84 ± 123.911 |
| | p-value | 0.1858 | 0.2127 | - |
| How full are you? | Baseline | 120.29 ± 102.665 | 132.93 ± 94.164 | 94.55 ± 113.482 |
| | Week 4 | 112.80 ± 102.799 | 130.66 ± 99.964 | 109.54 ± 116.441 |
| | p-value | 0.6146 | 0.9775 | - |
| How satiated are you? | Baseline | 97.08 ± 106.742 | 118.16 ± 102.146 | 94.29 ± 124.812 |
| | Week 4 | 93.24 ± 116.025 | 120.74 ± 102.263 | 113.82 ± 117.080 |
| | p-value | 0.3260 | 0.8309 | - |
| How strong is your desire to eat? | Baseline | -37.52 ± 102.824 | -52.10 ± 105.362 | -50.69 ± 97.813 |
| | Week 4 | -73.22 ± 113.570 | -81.53 ± 95.878 | -91.46 ± 120.316 |
| | p-value | 0.5589 | 0.6601 | - |
| How much do you think you could (or would want to) eat right now? | Baseline | -49.31 ± 99.192 | -55.93 ± 90.568 | -47.05 ± 92.694 |
| | Week 4 | -67.28 ± 114.521 | -78.41 ± 103.059 | -84.48 ± 102.535 |
| | p-value | 0.4052 | 0.6292 | - |

**Supplementary Table 2** Microbiota (family) abundance changes from baseline to weeks 4 and 12. Data are presented as Mean ± SD in the high-compliance population (n=100) and FAS population (n=125) for the 3.5g and 5g doses of arabinoxylan and placebo. Bolded values are significant ($p \leq 0.05$) within-group. * Significant $p \leq 0.05$, ** Significant $p \leq 0.01$, *** Significant $p < 0.001$ compared to placebo. Statistical significance markers are based on ANCOVA models adjusting for baseline, using least-squares means.

| | | High Compliance | | | FAS | | |
|---|---|---|---|---|---|---|---|
| **Family** | **Timepoint** | **3.5g (n=29)** | **5g (n=34)** | **Placebo (n=37)** | **3.5g (n=40)** | **5g (n=42)** | **Placebo (n=43)** |
| Bifidobacteriaceae | Baseline | 0.4422 ± 0.8581 | 0.5780 ± 0.8517 | 0.9317 ± 2.8752 | 0.4999 ± 0.8602 | 0.5423 ± 0.7820 | 0.8786 ± 2.7214 |
| | Week 4 | 0.6975 ± 1.0386 | 0.5587 ± 0.9196 | 0.5576 ± 1.3914 | 0.8165 ± 1.2087 | 0.5340 ± 0.8496 | 0.5315 ± 1.3513 |
| | Week 12 | 0.4817 ± 0.7825 | 0.5329 ± 1.0107 | 0.5517 ± 1.1580 | 0.4837 ± 0.7638 | 0.7004 ± 1.5190 | 0.5065 ± 1.1061 |
| Bacteroidaceae | Baseline | 35.6839 ± 24.6471 | 41.2054 ± 20.0414 | 41.1368 ± 18.9856 | 35.2259 ± 23.5087 | 38.6794 ± 20.7473 | 38.6687 ± 19.5734 |
| | Week 4 | 38.5199 ± 22.5138 | 39.9141 ± 24.1176 | 39.3580 ± 21.9233 | 37.9934 ± 22.8315 | 38.2693 ± 23.9805 | 37.4789 ± 22.5252 |
| | Week 12 | **46.4887 ± 23.3876** | 40.8995 ± 21.9544 | 44.6090 ± 22.9418 | 43.0203 ± 24.0570 | 38.7200 ± 23.5895 | 41.6961 ± 23.4804 |
| Lactobacillaceae | Baseline | 0.8773 ± 1.4453 | 0.1804 ± 0.4744 | 0.1721 ± 0.4905 | 0.6369 ± 1.2754 | 0.1506 ± 0.4348 | 0.1602 ± 0.4653 |
| | Week 4 | 0.3389 ± 0.7842 | 0.5373 ± 1.2443* | 0.2383 ± 0.6916 | 0.2483 ± 0.6773 | **0.4421 ± 1.1350*** | 0.2359 ± 0.6715 |
| | Week 12 | 0.3897 ± 0.8934 | 0.1744 ± 0.4063 | 0.1884 ± 0.5124 | 0.3102 ± 0.7862 | 0.2135 ± 0.4529 | 0.1771 ± 0.4873 |
| Ruminococcaceae | Baseline | 6.9261 ± 4.5103 | 9.7486 ± 6.0453 | 9.3348 ± 6.1602 | 8.4815 ± 6.2163 | 9.8755 ± 6.1280 | 9.4149 ± 6.1701 |
| | Week 4 | 6.9046 ± 4.9702 | 9.8096 ± 6.6485 | 9.5816 ± 7.1213 | 7.7734 ± 5.5909 | 10.1007 ± 6.3144 | 9.7125 ± 6.9369 |
| | Week 12 | 6.2974 ± 5.0521 | 9.2323 ± 4.9834 | 7.9345 ± 5.7842 | 7.1596 ± 5.9651 | 9.1803 ± 5.4708 | 8.6382 ± 6.3609 |
| Lachnospiraceae | Baseline | 19.5590 ± 12.6621 | 17.6065 ± 7.8316 | 17.8056 ± 8.2115 | 20.1096 ± 11.4743 | 18.4352 ± 7.7198 | 17.7954 ± 8.1151 |

| | | | | | | | |
|---|---|---|---|---|---|---|---|
| | Week 4 | 16.8099 ± 8.0001 | 18.6726 ± 13.0850 | 18.4324 ± 9.1870 | 17.0573 ± 7.4775 | 19.1611 ± 12.4939 | 18.7079 ± 9.0197 |
| | Week 12 | 16.3841 ± 9.3948 | 17.3534 ± 7.3765 | 17.2652 ± 7.5871 | 17.0070 ± 10.0014 | 18.0726 ± 7.8478 | 17.6357 ± 7.3017 |
| Akkermansiaceae | Baseline | 0.5944 ± 2.4271 | 0.0951 ± 0.2391 | 0.1847 ± 0.6845 | 0.5500 ± 2.0812 | 0.1489 ± 0.4669 | 0.1701 ± 0.6482 |
| | Week 4 | 0.1358 ± 0.3974 | 0.3651 ± 0.7101 | 0.1722 ± 0.4291 | 0.1855 ± 0.6007 | 0.3111 ± 0.6507 | 0.1617 ± 0.4176 |
| | Week 12 | 0.1640 ± 0.7896 | 0.2873 ± 1.1817 | 0.2589 ± 0.5738 | 0.2770 ± 0.8836 | 0.2460 ± 1.0781 | 0.5102 ± 1.7960 |

**Supplementary Table 3** Microbiota (genus) abundance changes from baseline to weeks 4 and 12. Data are presented as Mean ± SD in the high-compliance population (n=100) and FAS population (n=125) for the 3.5g and 5g doses of arabinoxylan and placebo. Bolded values are significant ($p \leq 0.05$) within-group. * Significant $p \leq 0.05$, ** Significant $p \leq 0.01$, *** Significant $p < 0.001$ compared to placebo. Statistical significance markers are based on ANCOVA models adjusting for baseline, using least-squares means.

| | | High Compliance | | | FAS | | |
|---|---|---|---|---|---|---|---|
| **Genus** | **Timepoint** | **3.5g (n=29)** | **5g (n=34)** | **Placebo (n=37)** | **3.5g (n=40)** | **5g (n=42)** | **Placebo (n=43)** |
| *Bacteroides* | Baseline | 2.6174 ± 4.6263 | 3.5343 ± 5.2809 | 4.7920 ± 8.9343 | 4.0753 ± 6.3718 | 3.9482 ± 5.4823 | 5.0023 ± 8.6011 |
| | Week 4 | 2.5259 ± 3.5380* | 2.8929 ± 4.1366** | **6.9656 ± 12.0580** | 3.8000 ± 6.1436 | 3.6779 ± 5.2103* | **6.8861 ± 11.6794** |
| | Week 12 | 2.2753 ± 3.0643 | 3.5336 ± 5.7106 | 5.5930 ± 9.6416 | 4.0251 ± 7.9151 | 4.1577 ± 5.9836 | 6.0022 ± 9.3627 |
| *Lactobacillus* | Baseline | 0.0083 ± 0.0332 | 0.0039 ± 0.0153 | 0.0000 ± 0.0000 | 0.0066 ± 0.0283 | 0.0032 ± 0.0139 | 0.0000 ± 0.0000 |
| | Week 4 | 0.0027 ± 0.0141 | 0.0127 ± 0.0643 | 0.0055 ± 0.0164 | 0.0019 ± 0.0120 | 0.0103 ± 0.0579 | 0.0052 ± 0.0159 |
| | Week 12 | 0.0014 ± 0.0073 | 0.0013 ± 0.0078 | 0.0009 ± 0.0038 | 0.0017 ± 0.0075 | 0.0031 ± 0.0143 | 0.0008 ± 0.0036 |
| *Akkermansia* | Baseline | 0.5944 ± 2.4271 | 0.0951 ± 0.2391 | 0.1847 ± 0.6845 | 0.5500 ± 2.0812 | 0.1489 ± 0.4669 | 0.1701 ± 0.6482 |
| | Week 4 | 0.1358 ± 0.3974 | 0.3651 ± 0.7101 | 0.1722 ± 0.4291 | 0.1855 ± 0.6007 | 0.3111 ± 0.6507 | 0.1617 ± 0.4176 |
| | Week 12 | 0.1640 ± 0.7896 | 0.2873 ± 1.1817 | 0.2589 ± 0.5738 | 0.2770 ± 0.8836 | 0.2460 ± 1.0781 | 0.5102 ± 1.7960 |
| *Prevotella* | Baseline | 33.0665 ± 27.3375 | 37.6711 ± 23.9234 | 36.3448 ± 23.3988 | 31.1506 ± 27.3707 | 34.7312 ± 24.7663 | 33.6664 ± 23.8450 |
| | Week 4 | 35.9940 ± 24.7425 | 37.0212 ± 27.0488 | 32.3924 ± 27.3254 | 34.1934 ± 26.0959 | 34.5914 ± 27.5397 | 30.5929 ± 27.4256 |
| | Week 12 | **44.2134 ± 25.9167** | 37.3659 ± 25.5266 | 39.0160 ± 28.2606 | 38.9952 ± 27.6604 | 34.5624 ± 27.5040 | 35.6939 ± 28.6857 |

**Supplementary Table 4** Postprandial iAUC of individual SCFAs and composite SCFA iAUC change from baseline to week 4. Data are presented as Mean ± SD including only subset participants in the high-compliance population (n=72) and FAS population (n=84) for the 3.5g and 5g doses of arabinoxylan and placebo. Bolded values are significant ($p \leq 0.05$) within-group. * Significant $p \leq 0.05$, ** Significant $p \leq 0.01$, *** Significant $p < 0.001$ compared to placebo. Statistical significance markers are based on ANCOVA models adjusting for baseline, using least-squares means.

| | | High Compliance | | | FAS | | |
|---|---|---|---|---|---|---|---|
| **SCFA** | **Timepoint** | **3.5g (n=20)** | **5g (n=25)** | **Placebo (n=27)** | **3.5g (n=27)** | **5g (n=29)** | **Placebo (n=28)** |
| Acetic Acid (h*uM) | Baseline | -25.2433 ± 35.0656 | -33.7323 ± 54.8677 | -37.8169 ± 55.1392 | -29.7831 ± 38.5777 | -35.0175 ± 55.0585 | -38.5568 ± 54.2499 |
| | Week 4 | -32.5836 ± 42.6432 | **-8.5304 ± 30.4224** | -27.2726 ± 42.8260 | -29.0123 ± 40.6373 | **-9.0325 ± 29.8580** | -26.1106 ± 42.4728 |
| Propanoic Acid (h*uM) | Baseline | 0.6774 ± 1.4280 | 0.0995 ± 2.0989 | 0.3101 ± 1.3377 | 0.1295 ± 1.8884 | 0.1503 ± 2.0390 | 0.2605 ± 1.3387 |
| | Week 4 | 0.4549 ± 1.9542 | 0.9026 ± 2.8610 | -0.0744 ± 1.7380 | 0.2368 ± 2.1052 | 0.8161 ± 2.6794 | 0.0923 ± 1.9115 |
| Butanoic Acid (h*uM) | Baseline | -0.2448 ± 0.9727 | -0.0921 ± 0.5322 | -0.1793 ± 0.7199 | -0.3556 ± 0.9536 | -0.1058 ± 0.5448 | -0.1755 ± 0.7062 |
| | Week 4 | -0.0376 ± 0.6209 | 0.1118 ± 0.6287 | -0.2875 ± 0.9327 | -0.1202 ± 0.6581 | 0.0389 ± 0.6426 | -0.2010 ± 1.0191 |
| Pentanoic Acid (h*uM) | Baseline | -5.1945 ± 10.0957 | -4.4962 ± 11.0830 | -4.7674 ± 9.8721 | -3.8394 ± 10.5138 | -4.8063 ± 10.5888 | -4.5769 ± 9.7213 |
| | Week 4 | -0.4073 ± 12.1170 | **0.8497 ± 12.8270** | 0.5727 ± 12.3581 | 0.0368 ± 11.4180 | **1.8575 ± 12.4772** | 0.5838 ± 12.0866 |
| Hexanoic Acid (h*uM) | Baseline | 0.1309 ± 1.6842 | 0.5419 ± 1.8240 | -1.0622 ± 4.8650 | 0.0567 ± 1.4562 | 0.6832 ± 2.0027 | -1.0482 ± 4.7746 |
| | Week 4 | 0.6044 ± 2.2094 | 0.7583 ± 3.3638 | 0.5563 ± 2.0318 | 0.4635 ± 1.9886 | 0.6422 ± 3.1307 | 0.5327 ± 1.9977 |
| Heptanoic Acid (h*uM) | Baseline | -4.0252 ± 7.9263 | -6.5764 ± 9.4245 | -4.2613 ± 15.2673 | -6.9958 ± 10.3060 | -6.4171 ± 8.8997 | -7.0682 ± 17.2176 |
| | Week 4 | -3.5459 ± 10.8657 | 0.9427 ± 7.2407 | -3.8032 ± 10.5326 | -3.0588 ± 10.8123 | 0.9427 ± 7.2407 | -3.8032 ± 10.5326 |

| | | | | | | | |
|---|---|---|---|---|---|---|---|
| Total (h*uM) | Baseline | -30.0038 ± 39.9193 | -34.6925 ± 60.5767 | -45.8068 ± 65.7485 | -34.8887 ± 44.4353 | -35.6612 ± 59.1707 | -47.4964 ± 65.1360 |
| | Week 4 | -36.5762 ± 46.7579 | **-2.0292 ± 36.5729** | -24.8127 ± 50.2072 | -30.7339 ± 45.5055 | -2.3620 ± 36.2163 | **-23.4812 ± 49.7699** |

**Supplementary Table 5** GI symptoms assessed by GSRS total and subscale score changes from baseline to weeks 4 and 12. Data are presented as Mean ± SD in the FAS population (n=125) for the 3.5g and 5g doses of arabinoxylan and placebo. Bolded values are significant ($p \le 0.05$) within-group. * Significant $p \le 0.05$, ** Significant $p \le 0.01$, *** Significant $p < 0.001$ compared to placebo. Statistical significance markers are based on ANCOVA models adjusting for baseline, using least-squares means.

| Parameter | Timepoint | 3.5g | 5g | Placebo |
|---|---|---|---|---|
| Abdominal Pain | Baseline | 1.2917 ± 0.5294 | 1.1905 ± 0.2958 | 1.1550 ± 0.2850 |
| | Week 4 | 1.2417 ± 0.5006 | 1.1905 ± 0.4364 | 1.2636 ± 0.4575 |
| | Week 12 | 1.1167 ± 0.2779 | 1.2033 ± 0.4142 | 1.2358 ± 0.5230 |
| Constipation | Baseline | 1.3167 ± 0.6957 | 1.2381 ± 0.5669 | 1.3256 ± 0.5704 |
| | Week 4 | 1.2000 ± 0.5433 | 1.2619 ± 0.4913 | 1.2016 ± 0.4492 |
| | Week 12 | 1.2500 ± 0.5736 | 1.1545 ± 0.3341 | 1.2927 ± 0.7424 |
| Diarrhea | Baseline | 1.0583 ± 0.1981 | 1.0635 ± 0.2679 | 1.1008 ± 0.3453 |
| | Week 4 | 1.1917 ± 0.4522 | 1.1508 ± 0.4368 | **1.3101 ± 0.6059** |
| | Week 12 | 1.1167 ± 0.2977 | 1.1301 ± 0.3864 | 1.1138 ± 0.2945 |
| Indigestion | Baseline | 1.350 ± 0.6883 | 1.280 ± 0.4788 | 1.419 ± 0.5281 |
| | Week 4 | 1.369 ± 0.7071 | 1.244 ± 0.4502 | 1.378 ± 0.5466 |
| | Week 12 | 1.250 ± 0.4082 | 1.268 ± 0.5012 | 1.311 ± 0.5695 |
| Reflux | Baseline | 1.20 ± 0.541 | 1.11 ± 0.391 | 1.12 ± 0.420 |
| | Week 4 | 1.14 ± 0.577 | 1.05 ± 0.185 | 1.16 ± 0.322 |
| | Week 12 | 1.16 ± 0.499 | 1.16 ± 0.361 | 1.17 ± 0.427 |
| Total | Baseline | 1.2433 ± 0.4620 | 1.1758 ± 0.2936 | 1.2233 ± 0.2918 |
| | Week 4 | 1.2279 ± 0.4422 | 1.1790 ± 0.2690 | 1.2632 ± 0.3173 |
| | Week 12 | 1.1792 ± 0.3201 | 1.1829 ± 0.2546 | 1.2248 ± 0.3897 |

.

**Supplementary Table 6** Mood assessed by BRUMS-24 domain score changes from baseline to weeks 4 and 12. Data are presented as Mean ± SD in the FAS population (n=125) for the 3.5g and 5g doses of arabinoxylan and placebo. Bolded values are significant ($p \leq 0.05$) within-group. * Significant $p \leq 0.05$, ** Significant $p \leq 0.01$, *** Significant $p < 0.001$ compared to placebo. Statistical significance markers are based on ANCOVA models adjusting for baseline, using least-squares means.

| Domain | Timepoint | 3.5g (n=40) | 5g (n=42) | Placebo (n=43) |
|---|---|---|---|---|
| Anger | Baseline | 0.1 ± 0.46 | 0.2 ± 0.58 | 0.4 ± 2.29 |
| | Week 4 | **0.0 ± 0.16** | **0.1 ± 0.37** | **0.1 ± 0.34** |
| | Week 12 | 0.2 ± 0.58 | **0.0 ± 0.00*** | 0.2 ± 0.91 |
| Confusion | Baseline | 0.2 ± 0.48 | 0.2 ± 0.61 | 0.4 ± 0.87 |
| | Week 4 | 0.2 ± 0.53 | 0.2 ± 0.66 | 0.2 ± 0.50 |
| | Week 12 | 0.2 ± 0.59 | 0.2 ± 0.77 | 0.4 ± 1.24 |
| Depression | Baseline | 0.2 ± 0.70 | 0.2 ± 0.58 | 0.4 ± 1.20 |
| | Week 4 | 0.4 ± 1.03 | 0.2 ± 0.70 | 0.4 ± 1.18 |
| | Week 12 | 0.3 ± 1.30 | 0.1 ± 0.69 | 0.2 ± 0.83 |
| Fatigue | Baseline | 1.7 ± 2.16 | 1.6 ± 2.06 | 2.3 ± 2.68 |
| | Week 4 | 2.3 ± 3.25 | 1.8 ± 2.51 | 2.3 ± 2.83 |
| | Week 12 | 1.7 ± 2.51 | 1.3 ± 1.76 | 2.1 ± 2.37 |
| Tension | Baseline | 0.6 ± 1.01 | 0.5 ± 1.29 | 0.8 ± 1.21 |
| | Week 4 | 0.6 ± 1.27 | 0.5 ± 1.11 | 0.6 ± 1.05 |
| | Week 12 | 0.4 ± 1.26 | 0.4 ± 1.34 | 0.4 ± 0.95 |
| Vigor | Baseline | 7.3 ± 3.11 | 7.5 ± 3.09 | 8.5 ± 4.27 |
| | Week 4 | 7.6 ± 3.47 | 8.1 ± 3.24 | 8.3 ± 4.20 |
| | Week 12 | **8.6 ± 3.70** | **9.3 ± 3.31**** | 8.3 ± 3.91 |

**Supplementary Table 7** HDL-C, high-density lipoprotein cholesterol; LDL-C, low-density lipoprotein cholesterol; TG, triglycerides. Blood lipid concentration and TG:HDL-C ratio changes from baseline to weeks 4 and 12. Data are presented as Mean ± SD in the FAS population (n=125) for the 3.5g and 5g doses of arabinoxylan and placebo. Bolded values are significant ($p \leq 0.05$) within-group. * Significant $p \leq 0.05$, ** Significant $p \leq 0.01$, *** Significant $p < 0.001$ compared to placebo. Statistical significance markers are based on ANCOVA models adjusting for baseline, using least-squares means

| Parameter | Timepoint | 3.5g (n=40) | 5g (n=42) | Placebo (n=43) |
|---|---|---|---|---|
| Total Cholesterol (mg/dL) | Baseline | 200.9 ± 45.91 | 194.2 ± 42.03 | 196.0 ± 38.85 |
| | Week 4 | 202.1 ± 62.02 | 187.4 ± 35.36 | 193.4 ± 44.94 |
| | Week 12 | **188.5 ± 33.69** | 188.8 ± 36.20 | 192.7 ± 36.61 |
| HDL-C (mg/dL) | Baseline | 53.9 ± 11.91 | 54.3 ± 17.21 | 57.8 ± 13.74 |
| | Week 4 | 52.6 ± 12.49** | 53.3 ± 13.02** | **63.3 ± 29.15** |
| | Week 12 | 54.8 ± 14.89 | 53.2 ± 15.43 | 57.2 ± 14.50 |
| LDL-C (mg/dL) | Baseline | 122.62 ± 41.728 | 116.78 ± 31.263 | 119.33 ± 31.374 |
| | Week 4 | 119.06 ± 44.072 | 112.19 ± 29.343 | 116.26 ± 32.905 |
| | Week 12 | **109.95 ± 29.995** | 113.39 ± 29.614 | 115.85 ± 34.293 |
| Triglycerides (mg/dL) | Baseline | 145.0 ± 94.74 | 124.8 ± 80.69 | 115.1 ± 70.47 |
| | Week 4 | 149.8 ± 127.56 | 116.1 ± 68.24 | 123.9 ± 81.82 |
| | Week 12 | 135.9 ± 81.31 | 129.5 ± 81.08 | 129.2 ± 131.37 |
| TG:HDL-C | Baseline | 2.9938 ± 2.3585 | 2.6971 ± 2.3150 | 2.2369 ± 1.8870 |
| | Week 4 | 3.2255 ± 3.4184 | 2.4515 ± 1.8654 | 2.3603 ± 2.3512 |
| | Week 12 | 2.7553 ± 2.0112* | 2.9614 ± 2.8044 | **2.7415 ± 4.0557** |

.

**Supplementary Table 8** HDL-C, high-density lipoprotein cholesterol; LDL-C, low-density lipoprotein cholesterol. Blood lipid concentrations adjusted by change in body weight changes from baseline to weeks 4 and 12. Data are presented as Mean ± SD in the high-compliance population (n=100) and FAS population (n=125) for the 3.5g and 5g doses of arabinoxylan and placebo. Bolded values are significant ($p \leq 0.05$) within-group. * Significant $p \leq 0.05$, ** Significant $p \leq 0.01$, *** Significant $p < 0.001$ compared to placebo. Statistical significance markers are based on ANCOVA models adjusting for baseline and body weight change, using least-squares means.

| | | **High Compliance** | | | **FAS** | | |
|---|---|---|---|---|---|---|---|
| **Parameter** | **Timepoint** | **3.5g (n=29)** | **5g (n=34)** | **Placebo (n=37)** | **3.5g (n=40)** | **5g (n=42)** | **Placebo (n=43)** |
| Total Cholesterol (mg/dL) | Baseline | 203.6 ± 46.58 | 195.7 ± 44.33 | 195.1 ± 38.34 | 200.9 ± 45.91 | 194.2 ± 42.03 | 196.0 ± 38.85 |
| | Week 4 | 200.4 ± 47.66 | **188.4 ± 34.11** | 194.3 ± 46.17 | 202.1 ± 62.02 | 187.4 ± 35.36 | 193.4 ± 44.94 |
| | Week 12 | **189.3 ± 35.73** | **189.7 ± 38.79** | 193.9 ± 37.25 | **188.5 ± 33.69** | 188.8 ± 36.20 | 192.7 ± 36.61 |
| HDL-C (mg/dL) | Baseline | 52.2 ± 12.02 | 53.8 ± 17.98 | 57.8 ± 14.43 | 53.9 ± 11.91 | 54.3 ± 17.21 | 57.8 ± 13.74 |
| | Week 4 | 52.2 ± 13.39* | 52.2 ± 12.41** | **64.8 ± 30.73** | 52.6 ± 12.49** | 53.3 ± 13.02** | **63.3 ± 29.15** |
| | Week 12 | 54.2 ± 16.80 | 53.3 ± 16.09 | 57.6 ± 15.03 | 54.8 ± 14.89 | 53.2 ± 15.43 | 57.2 ± 14.50 |
| LDL-C (mg/dL) | Baseline | 125.68 ± 44.891 | 117.97 ± 32.828 | 119.25 ± 31.484 | 122.62 ± 41.728 | 116.78 ± 31.263 | 119.33 ± 31.374 |
| | Week 4 | 122.74 ± 48.583 | 113.38 ± 26.644 | 117.38 ± 33.690 | 119.06 ± 44.072 | 112.19 ± 29.343 | 116.26 ± 32.905 |
| | Week 12 | **109.93 ± 31.121*** | 114.29 ± 31.843 | 116.78 ± 34.367 | **109.95 ± 29.995** | 113.39 ± 29.614 | 115.85 ± 34.293 |
| Triglycerides (mg/dL) | Baseline | 160.3 ± 98.93 | 131.2 ± 85.55 | 113.3 ± 68.60 | 145.0 ± 94.74 | 124.8 ± 80.69 | 115.1 ± 70.47 |
| | Week 4 | 172.5 ± 140.64 | 122.7 ± 72.32 | 120.2 ± 82.58 | 149.8 ± 127.56 | 116.1 ± 68.24 | 123.9 ± 81.82 |
| | Week 12 | 146.0 ± 81.13* | 130.4 ± 85.19 | 131.0 ± 137.70 | 135.9 ± 81.31 | 129.5 ± 81.08 | 129.2 ± 131.37 |

**Supplementary Table 9** Glucose and insulin AUC and Matsuda-derived index changes from baseline to week 4. Data are presented as Mean ± SD in the high-compliance (n=72) and FAS population (n=84) for the 3.5g and 5g doses of arabinoxylan and placebo. Bolded values are significant ($p \leq 0.05$) within-group. * Significant $p \leq 0.05$, ** Significant $p \leq 0.01$, *** Significant $p < 0.001$ compared to placebo. Statistical significance markers are based on ANCOVA models adjusting for baseline, using least-squares means.

| | | **High Compliance** | | | **FAS** | | |
|---|---|---|---|---|---|---|---|
| **Parameter** | **Timepoint** | **3.5g (n=20)** | **5g (n=25)** | **Placebo (n=27)** | **3.5g (n=27)** | **5g (n=29)** | **Placebo (n=28)** |
| Glucose (h*mg/dL) | Baseline | 408.700 ± 58.6545 | 406.732 ± 33.1824 | 406.333 ± 69.7615 | 404.36 ± 57.611 | 405.99 ± 38.958 | 405.95 ± 68.498 |
| | Week 4 | **431.838 ± 56.3643*** | 410.070 ± 40.2740 | 402.111 ± 63.1802 | 415.15 ± 58.454 | 399.97 ± 48.027 | 401.66 ± 62.044 |
| Insulin (h*mIU/L) | Baseline | 233.0488 ± 104.9491 | 249.3950 ± 114.8864 | 202.2472 ± 123.3218 | 198.60 ± 111.768 | 240.94 ± 117.577 | 199.36 ± 121.992 |
| | Week 4 | **277.3263 ± 191.8184** | 240.0500 ± 162.5668 | 196.8259 ± 95.39980 | **232.74 ± 183.938** | 221.46 ± 158.721 | 195.16 ± 94.042 |
| Matsuda-Derived Index | Baseline | 3.6101 ± 1.5675 | 3.8052 ± 1.9194 | 4.8182 ± 2.5671 | 4.7830 ± 2.8630 | 3.9702 ± 2.1249 | 4.8671 ± 2.5324 |
| | Week 4 | 3.5919 ± 2.1130 | 4.4354 ± 2.3417 | 5.5306 ± 4.1504 | 4.9015 ± 3.5658 | **5.1163 ± 3.3770** | 5.6233 ± 4.1023 |
